# Maternal Levels of Acute Phase Proteins in Early Pregnancy and Risk of Autism Spectrum Disorders in Offspring

**DOI:** 10.1101/2021.03.03.21252813

**Authors:** Martin Brynge, Renee M Gardner, Hugo Sjöqvist, Håkan Karlsson, Christina Dalman

## Abstract

Previous research supports a contribution of early-life immune disturbances in the etiology of autism spectrum disorders (ASD). Biomarker studies of the maternal innate (non-adaptive) immune status related to ASD risk have focused on one of the acute phase proteins (APP), C-reactive protein (CRP), with conflicting results. We evaluated levels of eight different APP in maternal serum samples drawn in first trimester, from 318 mothers to ASD-cases and 429 mothers to ASD-unaffected controls, nested within the register-based Stockholm Youth Cohort. Overall, we found no general trend of high levels of maternal APP being associated with increased risk of ASD. In contrast, maternal levels of CRP in the lowest compared to the middle tertile were associated with increased risk of ASD without ID or ADHD in offspring (OR = 2.15, 95 % CI 1.17-3.93). Further, levels of maternal ferritin in the lowest (OR = 1.82, 95 % CI 1.19-2.78) and highest (OR = 1.74, 95 % CI 1.16-2.60) tertiles were associated with increased risk of any ASD diagnosis in offspring, with stronger associations still between the lowest (OR = 3.58, 95 % CI 1.79-7.17) and highest (OR = 3.20, 95 % CI 1.62-6.29) tertiles of ferritin and risk of ASD with ID. The biological interpretation of lower CRP-levels among mothers to ASD-cases is not clear but might be related to the function of the maternal innate immune system. The finding of aberrant levels of ferritin conferring risk of ASD-phenotypes indicates a plausibly important role of iron during neurodevelopment.

## Introduction

The etiology of autism spectrum disorders (ASD) is complex, with plausible contributions of both genetic variation and early environmental exposures.^1^ ASD often co-occurs with other developmental disorders such as intellectual disability (ID) and attention deficit/hyperactivity disorder (ADHD)^2, 3^ defining diagnostic sub-groups with potentially different etiological pathways in terms of genetic and environmental influences.^4^

Evidence that immune-related proteins play pleiotropic roles during neurodevelopment and observations of immune anomalies among ASD-affected individuals^5, 6^ have led to the hypothesis that immune dysregulation during early life increases risk of ASD.^6-8^ Experimental animal models of maternal immune activation report autism-like phenotypes in the offspring, with decreased socialization and restricted patterns of behavior.^9^ However, the validity of such animal models for ASD remains to be established.^10^ Nevertheless, observational studies suggest that events associated with inflammation, such as maternal autoimmune disease, obesity and infections during pregnancy, are linked to ASD.^11-15^ The role of inflammation *per se* in the mechanisms underlying these associations is uncertain, and the observed associations may also partly be explained by genetic confounding.^16-18^

To understand if the maternal immune activation hypothesis is relevant for ASD, numerous studies have explored maternal immune-related biomarkers from blood samples collected during pregnancy. These studies have mostly focused on cytokines, with inconsistent results.^19-21^ Cytokines are powerful regulators of the immune system that also play roles in CNS development, but quantitation is technically challenging due to generally low baseline levels with transient secretion patterns and short half-lives. The acute phase proteins (APP) are involved in the initial non-adaptive (innate) immune response,^22^ although several APP exhibit additional functions, not apparently related to immune function.^23-25^ They generally have longer half-lives compared to cytokines and base-line concentrations in the measurable range. The few previous studies on gestational APP and risk of ASD, measured only one of the APP, C-reactive protein (CRP),^26-29^ with inconsistent results.

In the current study, we measured eight different APP in sera collected in the first trimester of pregnancy and estimated the risk of ASD in the offspring associated with the individual APP, using a large, well-characterized population-based cohort.

## Materials and Methods

### Study Population

The present study employed a case-cohort study design nested within the population-based register linkage Stockholm Youth Cohort (SYC).^30^ The source population consisted of children born 1996-2000 (n=98 597) (Supplementary Figure 1). In Stockholm County, maternal serum samples obtained during the national screening program of pregnant women have been stored frozen since 1998. We retrieved 0.2 ml serum from all available maternal sera samples, for ASD-cases (n=430) and mothers to control individuals (n=549), from those individuals who also had a neonatal dried blood spots (NDBS) as described in detail previously.^31^ Samples were obtained at median gestational week 10.6 (interquartile range: 9.3 – 12.5 weeks) representing a period near the end of the first trimester.

For the analysis in this study, we included only those samples drawn in the first trimester (≤13 weeks), since maternal serum proteins may vary by gestational age due to maternal blood volume expansion and altered metabolism.^32, 33^

Our final study sample consisted of 318 mothers to ASD cases and 429 mothers to control individuals. Mothers to individuals in the final study sample were generally similar to the source population, though they tended to be older, have higher educational level, be of higher socioeconomic status, and were less likely to be born in Asia or Africa (Supplementary Table 1). Ethical approval was obtained by the Stockholm regional review board (DNR 2011/695-31/2, amendment 2012/706-32). Individual consent was not required for this anonymized register-based study.

### Case Ascertainment

The case-finding procedure has been described previously.^30, 31^ We considered any ASD diagnosis as an outcome (regardless of comorbidities) and also stratified the outcomes as ASD only (without comorbid ID or ADHD), ASD with ID, and ASD with ADHD. Individuals with both comorbid ID and ADHD were included in the ASD with ID group.

### Laboratory Analysis

Serum aliquots were stored at −80°C until analysis. After thawing on ice, samples were diluted 1:10,000 for analysis of α-2-macroglobulin (A2M), haptoglobin (HAP), CRP and serum amyloid P (SAP), or 1:100 for analysis of ferritin (FER), fibrinogen (FIB), procalcitonin (PCT), serum amyloid A (SAA) and tissue plasminogen activator (tPA). Samples from cases and controls were assigned randomly to 96-well assay plates for analysis using commercially available premixed, multiplex panels and the Bio-Plex 200 System (Bio-Rad, Hercules, CA, USA).

The average coefficient of variation for manufacture-provided control samples over the 13 assay plates was 18.6% (Supplementary Table 2). Values below the lower limit of quantitation (LLOQ) were assigned a value of LLOQ/√2. Values above the upper limit of quantitation (ULOQ) were assigned a value of ULOQ×1.1. HAP had a large proportion of imputed values (75.77%), with the majority above the ULOQ, and was therefore excluded from the final analysis.

### Covariates

Covariates considered as potential confounders were chosen on the basis of previously described relationships with ASD and plausibility of a potential relationship to APP: maternal age, psychiatric history, BMI, region of birth, education, and smoking at first antenatal visit; fetal sex; birth order; family income; and gestational week and season at serum sample. Covariates were extracted from the Medical Birth Register and the National Patient Register. Sociodemographic data were extracted from the Integrated Database for Labor Market Research.

### Statistical Analysis

The concentrations of APP were log-transformed to normalize the distributions (Supplementary Figure 2A). To reduce the influence of assay plate-to-plate variation, we calculated standardized plate-based z-scores by subtracting the plate-specific mean concentration from each measurement and dividing by the plate-specific standard deviation (Supplementary Figure 2B). The standardized scores were then categorized into tertiles based on the distribution among controls (Supplementary Figure 2C).

Covariates were tested for association with the z-scores of each APP among cases and controls separately. We conducted linear regression analyses with the covariates as the independent variable and each APP as outcome variables, followed by a joint Wald-test to examine if the categories of the covariates were associated with different mean levels of APP. Covariates were included in the adjusted models if even weakly associated (p<0.2) with at least one of the APP among the randomly-sampled control population and with any of the outcomes. The final adjusted model included sex; birth order; family income quintile; maternal BMI, psychiatric history, region of origin, and age.

In the categorical analyses, we used logistic regression models to estimate the odds of ASD associated with the different APP, using the middle tertile as the referent and followed by a Wald-test for the association between each APP and odds of ASD. For continuous analyses, we used restricted cubic spline models with three knots and a z-score=0 as the referent. To examine the overall association each APP with odds of ASD, we tested if all spline terms jointly were equal to zero using the Wald test. We investigated evidence for potential non-linearity by testing if all spline terms that would indicate a change in the direction of the relationship were equal to zero.

We conducted sensitivity analyses by restricting the cohort to the sample of Nordic-born mothers, as maternal region of origin was strongly associated with some APP and also with the likelihood of a child receiving an ASD with ID diagnosis. The non-Nordic group (n=81) was too small to evaluate on its own. In a separate sensitivity analysis, we adjusted additionally for annual quarter (season) at serum sampling and gestational week at serum sampling, factors that were not associated with ASD, but with plausible influence on APP levels.

## Results

### Association of covariates with case status

As expected, ASD-cases were more likely to be male and firstborn (Table 1). Mothers to ASD-cases were more likely to be older, have a history of psychiatric illness, and have lower family income levels, and were less likely to have a normal BMI, compared with mothers to unaffected controls. Mothers born in Africa and Asia were overrepresented among children with ASD and co-morbid ID.

**Table 1.**
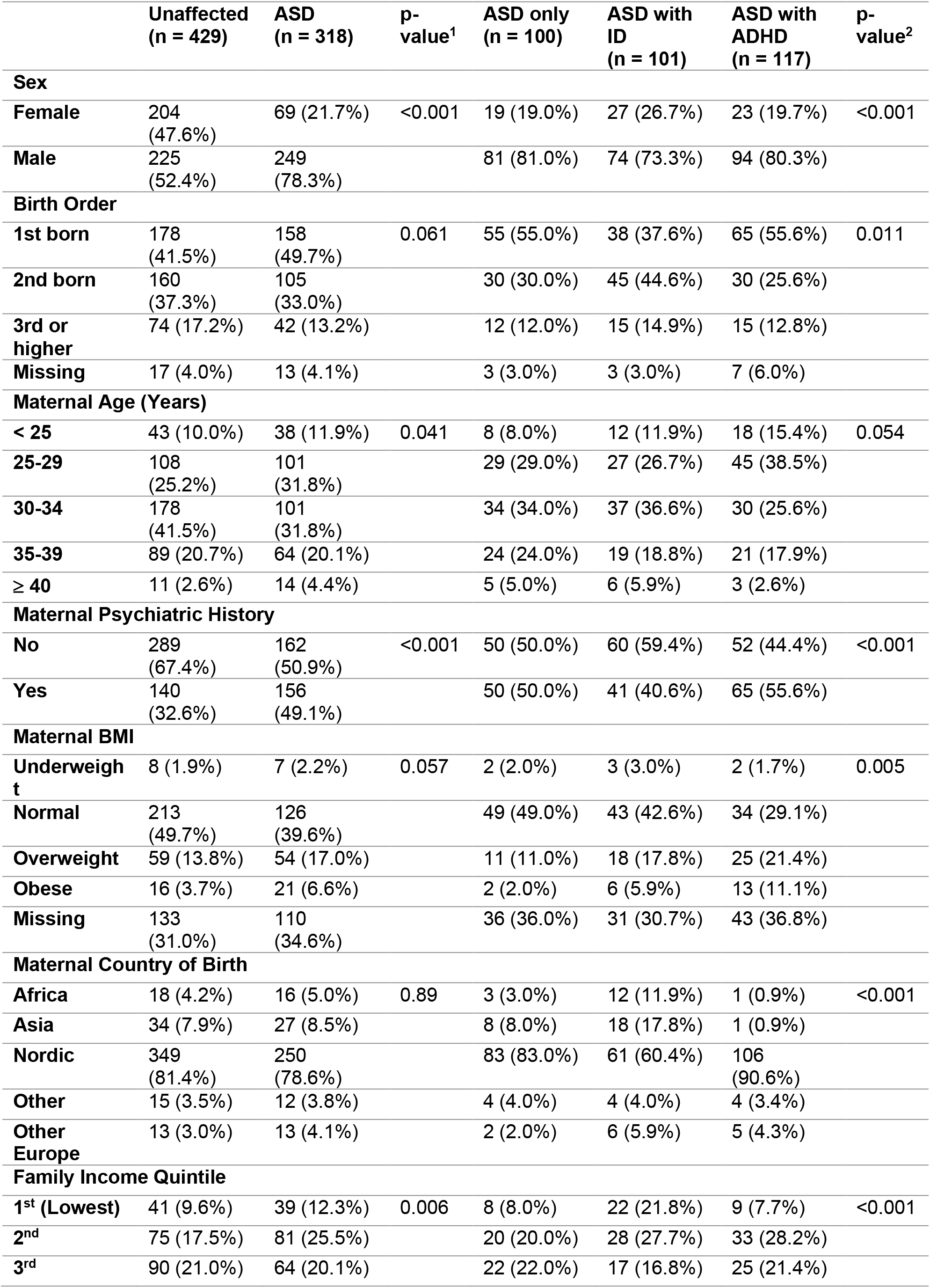
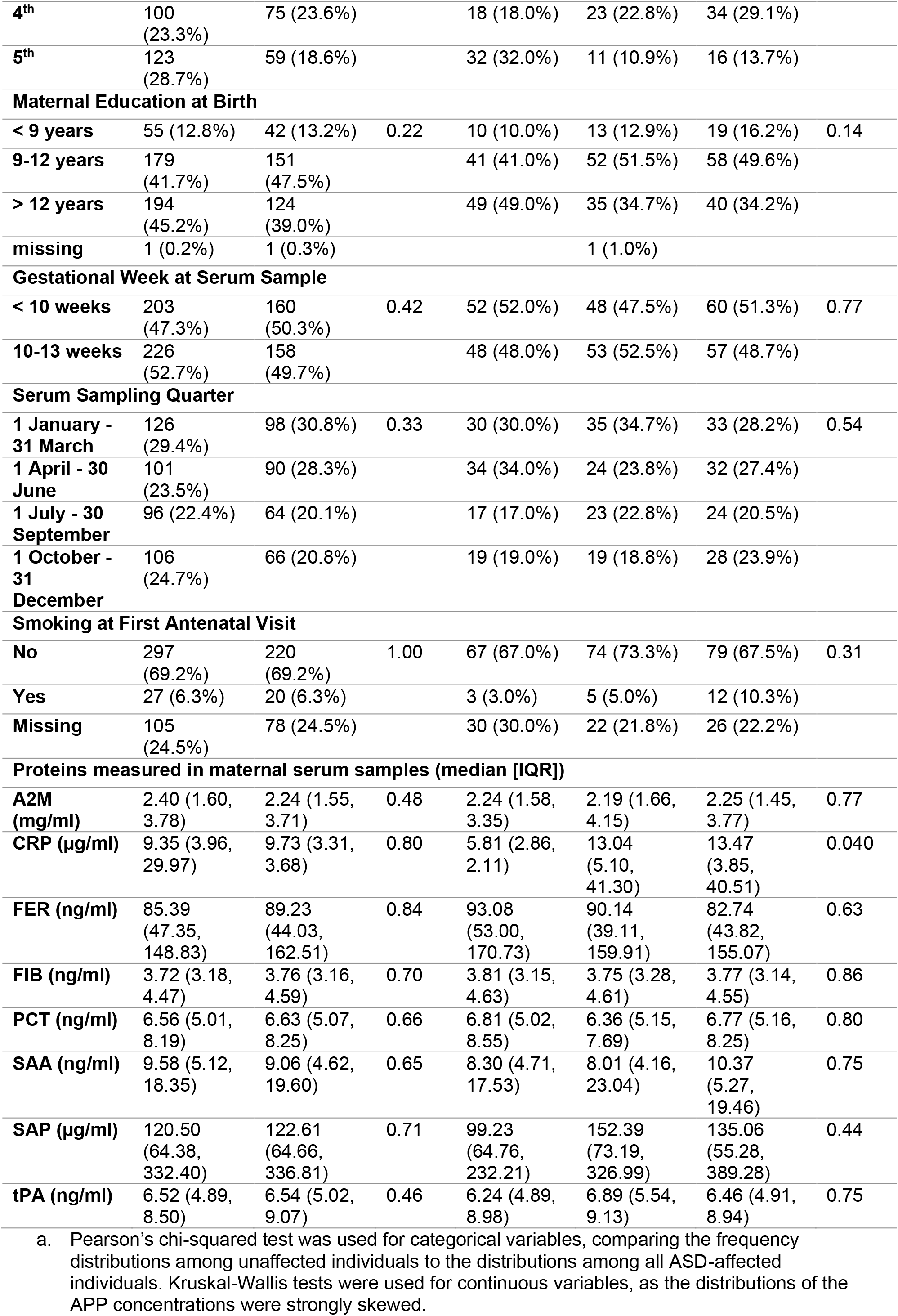
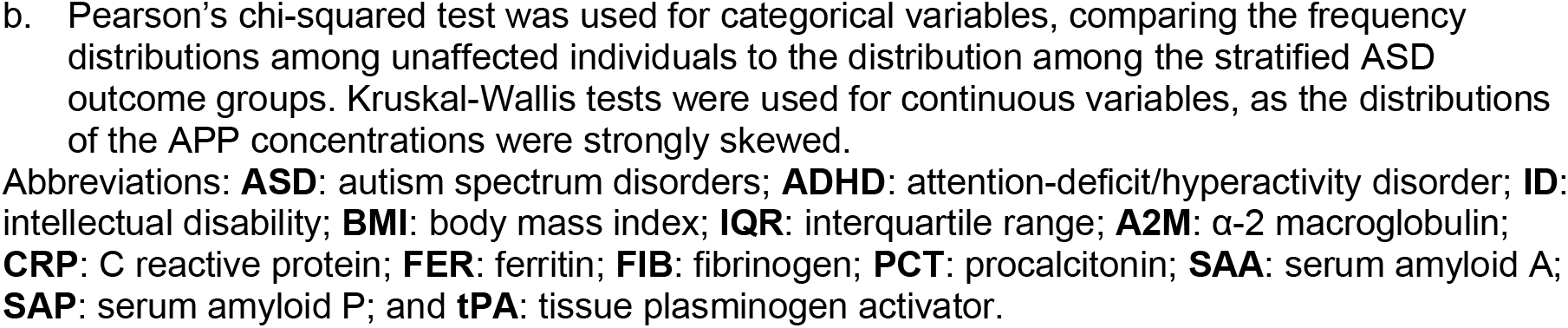
Characteristics of individuals diagnosed with ASD and unaffected individuals in the study sample.

### Association of APP with each other and with covariates

Among mothers to unaffected controls, 14 of the 28 possible pairwise combinations of maternal APP (Supplementary Figure 3) were positively correlated at a significance level p<0.05, with the strongest correlations observed between CRP and A2M; CRP and SAP; A2M and SAP; and tPA and PCT.

Associations (p<0.2) with at least one APP were seen for all covariates except serum sampling quarter in mothers to unaffected individuals (Figure 1, Supplementary Table 3). Maternal BMI was among the strongest predictors of APP levels. Similarly, among mothers to ASD-affected individuals, all covariates were associated (p<0.1) with at least one APP, except sex and maternal age (Supplementary Figure 4, Supplementary Table 4).

**Figure 1.**
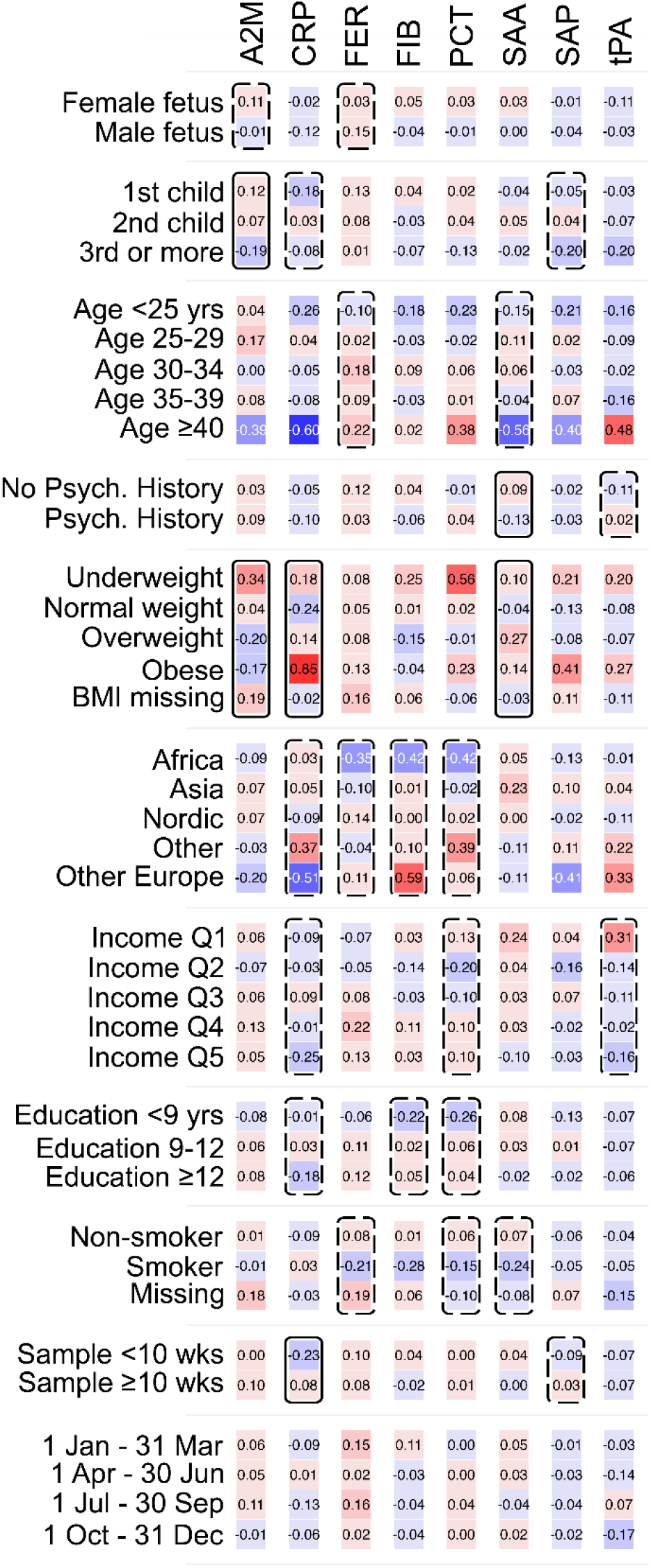
Heat map showing the mean APP z-score by categories of the covariates, among 429 unaffected individuals in the cohort. Solid boxes indicate that the APP is associated with the covariate at p<0.05. Dashed boxes indicate that the APP is associated with the covariate at p<0.20. Abbreviations: **A2M**: α-2 macroglobulin; **CRP**: C-reactive protein; **FER**: ferritin; **FIB**: fibrinogen; **PCT**: procalcitonin; **SAA**: serum amyloid A; **SAP**: serum amyloid P; **tPA**: tissue plasminogen activator; **Psych:** Psychiatric; **BMI**: body mass index; **Income Q**: income quintile.

### Association of APP with odds of ASD

Compared with ASD-unaffected controls, there were no significant differences in median levels of APP in mothers to ASD-affected individuals, though there was a reduction (p<0.05) in CRP among mothers to offspring with ASD only (Table1, Supplementary Figure 5).

In the unadjusted regression analysis of APP tertiles, we observed a trend towards a u-shaped association (p=0.06) between FER and odds of any ASD (Supplementary Figure 6), with a similar u-shaped pattern present for the stratified outcome ASD with ID (p<0.01) but absent for the other stratified outcomes. The lowest tertile of CRP was associated with increased odds of ASD only (p<0.01).

In fully adjusted models, the association between FER and any ASD was strengthened with increased odds of ASD in the lowest (OR = 1.82, 95 % CI 1.19-2.78) and highest (1.74, 1.16-2.60) tertiles compared to the middle tertile (Figure 2). The association between FER and odds of ASD with ID was somewhat attenuated, with increased odds of ASD with ID for the lowest (3.58, 1.79-7.17) and highest (3.20, 1.62-6.29) tertiles. The association between CRP and odds of ASD only was attenuated, although with increased odds for the lowest tertile compared to the middle tertile (2.15, 1.17-3.93).

**Figure 2.**
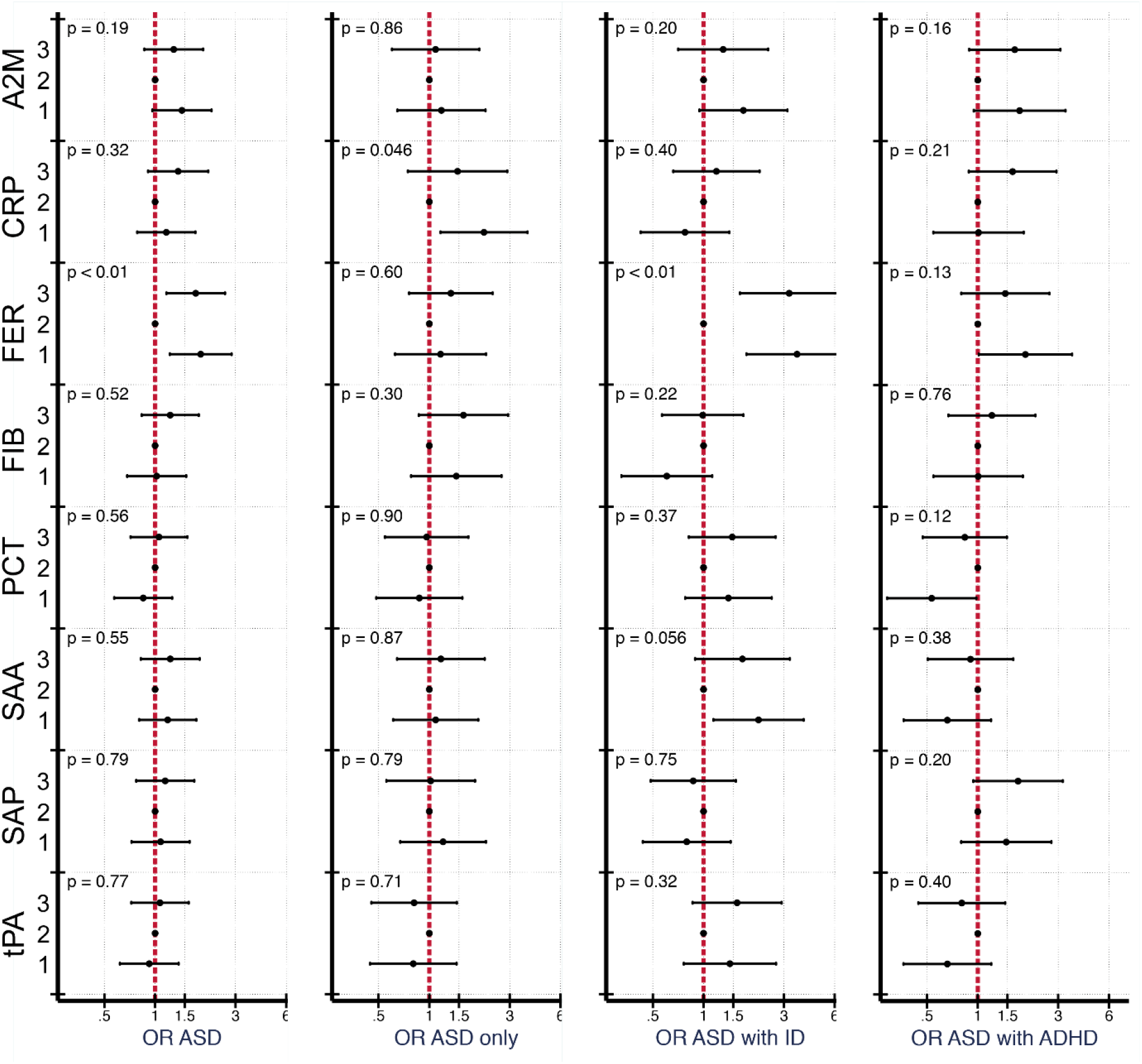
The relationship between APP and odds of ASD, stratified by co-occurrence of ID and ADHD, when comparing 318 ASD cases to 429 unaffected individuals selected from the cohort. Tertiles of each APP were created using the distribution of z-scores among unaffected individuals to set the cut-offs and the middle quintile was used as the referent category. Models were adjusted for sex, birth order, maternal BMI, maternal psychiatric history, maternal region of origin, maternal age and family income quintile. P-values are shown for a Wald test with a null hypothesis that all APP categorical terms were jointly equal to zero, as a test of whether each APP was generally associated with the outcome. Abbreviations: **A2M**: α-2 macroglobulin; **CRP**: C-reactive protein; **FER**: ferritin; **FIB**: fibrinogen; **PCT**: procalcitonin; **SAA**: serum amyloid A; **SAP**: serum amyloid P; and **tPA**: tissue plasminogen activator.

In adjusted cubic spline models, we observed evidence for non-linear relationships between z-scores of maternal CRP, FER and SAA and the outcomes (Supplementary Table 5). As in the categorical models, we observed a u-shaped relationship between FER and any ASD (p=0.08) as well as ASD with ID (p=0.05; Figure 3, Supplementary Figure 7). The overall pattern of association between CRP and odds of ASD only, with higher odds at lower concentrations, was similar to the categorical analysis, although it did not reach statistical significance (p=0.35) (Supplementary Figure 8). A u-shaped pattern of association was observed between maternal CRP (p=0.06) and SAP (p=0.03) and odds of ASD with ADHD (Supplementary Figure 9), with the strongest associations above the mean.

**Figure 3.**
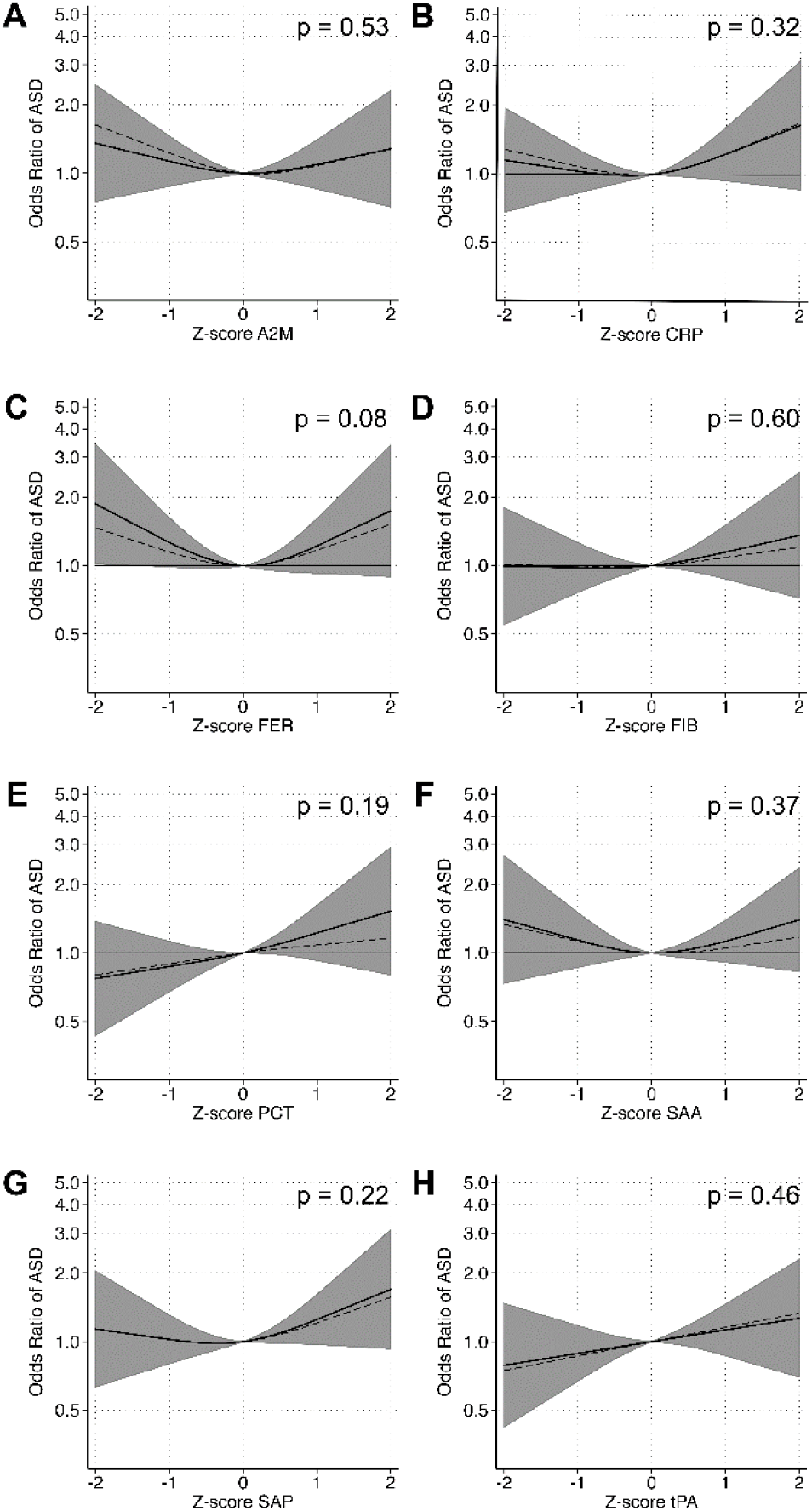
The relationship between APP and odds of ASD when comparing 318 individuals affected by ASD to 429 unaffected individuals selected from the cohort. Each panel displays the odds of ASD according to APP z-score, flexibly fit using restricted cubic spline models with three knots and a z-score=0 as the referent. The dashed line represents the unadjusted estimate of the relationship between each APP and odds of ASD. The solid line represents the fully adjusted model, adjusted for sex, birth order, maternal BMI, maternal psychiatric history, maternal region of origin, maternal age and family income quintile. The gray bands represent the 95% confidence interval for the fully adjusted model. P-values are shown for a Wald test with a null hypothesis that all APP spline terms were jointly equal to zero, as a test of whether each APP was generally associated with the outcome. Abbreviations: **A2M**: α-2 macroglobulin; **CRP**: C-reactive protein; **FER**: ferritin; **FIB**: fibrinogen; **PCT**: procalcitonin; **SAA**: serum amyloid A; **SAP**: serum amyloid P; and **tPA**: tissue plasminogen activator.

### Sensitivity analyses

The estimates observed in the sensitivity analyses, either restricting to Nordic-born mothers or considering factors that might influence APP-levels, were similar to those observed in the main analyses (Supplementary Figure 10, Supplementary Figure 11).

## Discussion

In this population-based cohort of pregnant women, we measured a range of immune markers to assess the possible link between innate immune activation in early pregnancy and subsequent ASD in offspring. Overall, we found no strong evidence for elevated APP being associated with increased odds of ASD regardless of comorbidities. However, we did observe that low levels of CRP were associated with increased odds of ASD without ID or ADHD, with some analyses suggesting that higher levels of CRP and SAP are associated with higher risk of ASD with ADHD. We also found that both low and high levels of FER were associated with increased odds of ASD, particularly ASD with ID.

### Comparison with previous studies

There are four previous studies investigating maternal APP and risk of ASD in offspring, all measuring the acute phase reactant CRP.

In a study by Brown et. al,^26^ the highest quintile of gestational CRP in first and early second trimester was associated with increased odds for autism, and a positive association was apparent when CRP was treated as a continuous variable. The prevalence of ASD in that study (1,132 cases of childhood autism in a birth cohort of 1.2 million, or 0.094%) stands in contrast to the present study, where we included all ICD and DSM-IV autistic diagnoses, with a joint prevalence of 1.52%. Furthermore, we were able to control for a larger number of health- and sociodemographic covariates, such as BMI and socioeconomic status, which may also contribute to the discrepancy in the results.

In a recent study by Egorova *et al*., 62 biomarkers including CRP were measured in serum samples (gestational week 14) of 100 mothers to ASD cases and 100 unaffected controls.^29^ No significant associations with CRP were found, though this smaller study had decreased power to detect differences. Also, CRP was treated as a linear continuous variable, in contrast to the present study where we allowed for non-linear relationships.

Koks *et al*. reported an association between maternal CRP (measured before 18 weeks of gestation) and autistic traits in offspring at age 6, as measured by the continuous Social Responsiveness Scale in the Generation R cohort study.^28^ The association was completely attenuated after controlling for maternal health-related factors and socioeconomic covariates, stressing the importance of controlling for such parameters.

In the Californian Early Markers for Autism (EMA) case-control study, the third and fourth quartiles of CRP (at 15-19 weeks of gestation) compared to the lowest quartile were associated with a decreased risk of ASD,^27^ which is in general agreement with our current findings. The definition and prevalence of the outcome were similar, as were the regression models with respect to covariates included.

### Interpretation and potential mechanisms

#### Maternal APP and risk for ASD

Baseline concentrations of APP are influenced by both genetic and environmental factors. Previous studies have estimated heritability coefficients in adults for FIB (27-51%), tPA (23%-27%), and CRP (10-65%), with estimates from a longitudinal twin study at around 50% for CRP, with stability over time.^34-36^

We observed evidence of moderate to high pairwise correlations between several of the APP, possibly reflecting shared regulation. However, many of the observed relationships were weak or non-existent, implying partially independent modes of regulation or biological function, in line with previous knowledge of the APP.^22^ In general, lower correlations were observed among APPs in maternal serum compared to those observed in neonatal samples.^31^ In accordance with previous evidence,^37^ we observed a strong correlation between maternal BMI and CRP, with overweight and obesity being associated with elevated levels.

Several immune proteins have dual roles, including a role as modulators of neurodevelopment. Animal models and observational studies suggest a harmful effect of maternal infections during pregnancy on the developing fetal brain, not only of pathogenic (teratogenic) microorganisms themselves, but also an effect of maternal inflammatory mediators crossing the fetal-placental barrier.^9, 10^ This is often referred to as the maternal immune activation hypothesis of ASD. Based on the eight APP studied here, we found no convincing general trend of maternal immune activation early in ASD pregnancies. On the contrary, low levels of CRP were associated with increased odds of ASD without ID or ADHD. We also observed a borderline association between increased odds of ASD with ID and low levels of SAA, an APP with secretion kinetics resembling those of CRP.^38^

#### CRP and ASD

CRP is a member of the pentraxin family, synthesized in hepatocytes as a response to proinflammatory cytokines, particularly Interleukin-6, and to a lesser degree Interleukin-1 and Tumor Necrosis Factor-α.^39^ It opsonizes common molecular patterns on the surface of pathogens and induces complement activation.^36^

Evidence of an association between low levels of maternal CRP and increased odds of ASD are now apparent in two large case-control/cohort studies of similar design.^27^ In schizophrenia, another complex neuropsychiatric disorder sharing common genetic variation, and brain transcriptional dysregulation with autism,^40, 41^ genetic loci associated with high levels of CRP have been found protective in Mendelian randomization studies.^42^ This is in alignment with a previous observational study of several neonatal APP and risk of non-affective psychotic disorders in adulthood.^43^ In our case-cohort study of neonatal APP measured in dried blood spots, we observed a u-shaped association between CRP and odds of ASD.^31^

It is not clear how low levels of maternal CRP in first trimester might influence the risk of ASD in offspring. Considering the opsonizing and complement activating functions of CRP, low levels might represent a suboptimal innate immune effector function. Further, low levels of CRP in the first trimester do not exclude maternal or fetal immune activation later in pregnancy, and the low concentrations early in pregnancy might influence risk of serious infections later on.^31^

#### Ferritin and ASD

Serum FER is a sensitive marker of total body iron not bound to hemoglobin.^44^ There is increasing evidence concerning the importance of iron metabolism for early neurodevelopment.^45^ Apart from serving as oxygen transporter, iron functions as a cofactor in cytochrome reactions generating ATP and in enzymes involved in neuronal myelinization.^46-48^ In an observational study, Schmidt *et al*. reported an association between self-reported low maternal iron intake prior to and during pregnancy and risk of autism in the offspring.^49^ Maternal iron deficiency is the most common cause of anemia, and the latter is associated with several adverse birth-related and behavioral outcomes in the offspring, including small for gestational age, ASD, ADHD and ID.^14, 50, 51^ Low FER levels in neonates have been associated with adverse neurocognitive and behavioral outcomes.^52-54^ In our previous case-cohort study of neonatal APP, we observed an increased risk of ASD with lower levels of FER if mothers had anemia during gestation.^31^ In a separate discordant sibling-comparison, levels of FER below the mean were strongly associated with increased odds of ASD.

While low levels of FER are specific for iron deficiency, high levels are harder to interpret, since they can be a consequence iron overload or elevated as part of the acute phase response.^23^ If a consequence of iron-overload, high levels of FER might indicate increased levels of circulating non-bound iron that may in turn lead to oxidative stress and DNA damage and affect the maternal-placental-fetal unit.^55, 56^ There is evidence of u-shaped associations between maternal hemoglobin and FER levels with obstetric outcomes, such as low birth weight.^51^ The increase in FER may also represent increased innate immune activity. The hypoferraemia due to increased concentrations of iron chaperones, such as FER, induced by inflammation is important in the host defense against infection by sequestering iron from pathogens.^57^ However, we did not detect any risk associated with high levels of any of the other APP studied to indicate an ongoing increase in innate immune activity.

#### Strengths and weaknesses

By measuring a range of proteins involved in the maternal innate immune defense in a large population-based cohort, we aimed to increase the likelihood of picking up any signal of maternal immune activation. Further, due to the pleiotropic physiological roles of most APP, we also had the possibility to investigate other pathophysiological processes, not necessarily directly related to immune functioning. We used a validated case-finding procedure^30^ and a broad range of register data to control for health- and sociodemographic confounders.

Although the sample size is large, it may be underpowered to detect subtle associations, and the number of individuals included in our analyses of the stratified outcomes was particularly limited. Further, although it can be considered a strength to include many immune markers, we also make multiple statistical comparisons, increasing the probability of chance findings. We only have a single measurement at one time-point in pregnancy, restricted to samples drawn in the first trimester for the purpose of interpretability. Consequently, we cannot draw conclusions regarding these markers in later trimesters. By restricting to samples drawn in the first trimester, we excluded a larger proportion of migrant women and women from low-income families. However, a separate sensitivity analysis stratifying the sample on maternal region of birth did not change the estimates. Further, since this is an observational study, we cannot give the results a causal interpretation. Although we adjust for a large number of covariates, there is still the possibility the results are influenced by residual confounding. Finally, the coefficient of variation was markedly higher for PCT compared to the other analytes, and thus the results for PCT must be interpreted with caution.

#### Conclusion

We investigated associations between eight maternal inflammatory proteins in first trimester of pregnancy and risk of ASD in offspring. We observed increased odds of ASD without ID or ADHD in offspring of mothers with low levels of CRP, though the biological mechanism underlying this relationship remains unclear. We also observed increased odds of any ASD, and particularly ASD with ID, in offspring of mothers with both low and elevated levels of ferritin. Our results give no strong support for the maternal immune activation hypothesis, but rather suggest that low maternal innate immune activity is associated with adverse neurodevelopmental outcomes. Our results also add evidence to a plausibly important role of iron metabolism during early neurodevelopment. Future studies ought to investigate the causes of low CRP levels in early pregnancy and how these relate to ASD risk in the offspring, and consider measurements of additional biomarkers of iron status to improve our understanding of the relationship between FER and ASD/ID.

## Data Availability

The Swedish health and population register data used in this study are available from Statistics Sweden and the Swedish National Board of Health and Welfare. The authors are not allowed to distribute the data according to the ethical approval for this study and the agreements with Statistics Sweden and the Swedish National Board of Health and Welfare.

## Acknowledgements

This work was supported by grants from the Swedish Research Council (grant numbers 2016-01477, 2012-2264, 523-2010-1052 [to CD], grant number 2017-02900 [to RG]) and the Stanley Medical Research Institute (to HK). The funders had no role in the design and conduct of the study; collection, management, analysis, and interpretation of the data; preparation, review, or approval of the manuscript; or decision to submit the manuscript for publication.

