## Supplementary Information for "Maternal Levels of Acute Phase Proteins in Early Pregnancy and Risk of Autism Spectrum Disorders in Offspring"

**This files includes Supplementary Figures 1-11, Supplementary Tables 1-5, STROBE checklist, and ICMJE disclosure forms**

**<u>Supplementary Figures:</u>**

**Supplementary Figure 1.** Selection of individuals with maternal serum samples.

**Supplementary Figure 2.** Levels of one maternal acute phase protein (ferritin) according to analytic plate.

**Supplementary Figure 3.** The Spearman rho correlations of eight maternal acute phase proteins with each other, as measured in maternal serum samples of 429 unaffected controls selected from the cohort.

**Supplementary Figure 4.** Heat map showing the mean maternal acute phase protein z-score according to each category of the covariates, among 318 ASD-affected individuals in the cohort.

**Supplementary Figure 5.** The distribution of each acute phase protein measured in maternal serum samples, by ASD-case status.

**Supplementary Figure 6.** The unadjusted relationship between maternal acute phase proteins and odds of ASD, stratified by co-occurrence of ID and ADHD, when comparing 318 ASD cases to 429 unaffected individuals selected from the cohort.

**Supplementary Figure 7.** The relationship between maternal acute phase proteins and odds of ASD with co-occurring ID when comparing 101 individuals affected by ASD with co-occurring ID to 429 unaffected individuals selected from the cohort.

**Supplementary Figure 8.** The relationship between maternal acute phase proteins and odds of ASD without co-occurring ID or ADHD when comparing 100 individuals affected by ASD with co-occurring ID to 429 unaffected individuals selected from the cohort.

**Supplementary Figure 9.** The relationship between maternal acute phase proteins and odds of ASD with co-occurring ADHD when comparing 117 individuals affected by ASD with co-occurring ID to 429 unaffected individuals selected from the cohort.

**Supplementary Figure 10.** The relationship between maternal acute phase proteins and odds of ASD in a sensitivity analysis where the study sample is restricted to Nordic-born mothers.

**Supplementary Figure 11.** The relationship between maternal acute phase proteins and odds of ASD in a sensitivity analysis where covariates related to the timing of sampling were included in the model.

**<u>Supplementary Tables:</u>**

**Supplementary Table 1.** Characteristics of individuals in the source population, those for whom neonatal dried blood spots and maternal serum samples were selected, and those for whom samples were selected for acute phase protein analysis.

**Supplementary Table 2.** Quality control statistics for multiplex assays to analyze acute phase protein concentrations in maternal serum samples.

**Supplementary Table 3.** P-values for the association of maternal acute phase proteins with other covariates among mothers to 429 unaffected individuals.

**Supplementary Table 4.** P-values for the association of maternal acute phase proteins with other covariates among mothers to 318 ASD-affected individuals.

**Supplementary Table 5.** P-values for non-linearity in restricted cubic spline analyses.

**Supplementary Figure 1.** Selection of individuals with maternal serum samples from the Stockholm Youth Cohort (SYC) source population.

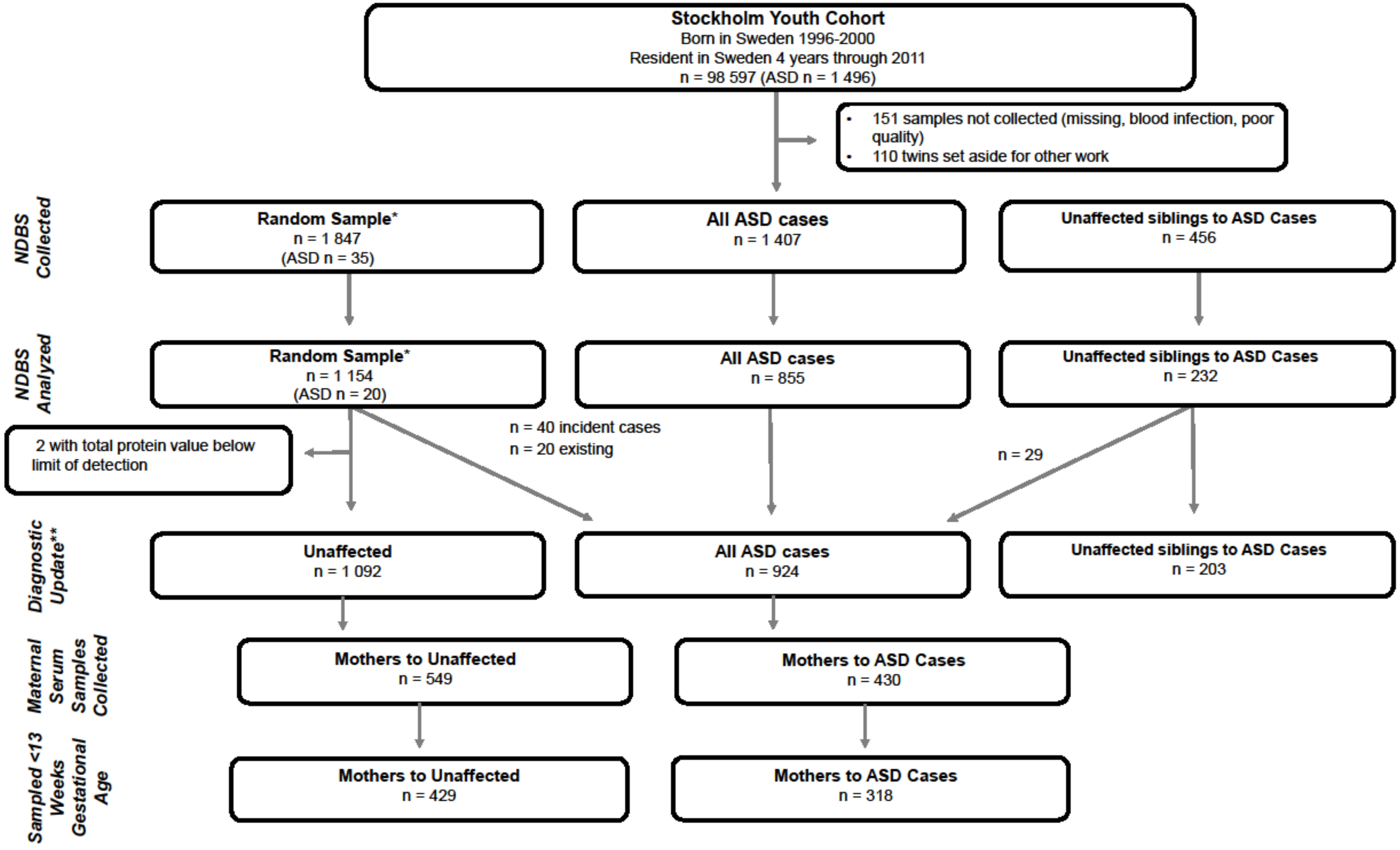

*A random sample of SYC that includes individuals with ASD also included among the total ASD cases sampled.

** Diagnostic update as of 2016-12-31, that resulted in 29 incident cases among unaffected siblings to ASD cases, and 40 incident cases among the ASD-unaffected control individuals.

**Supplementary Figure 2.** Boxplots of log-2-values of Ferritin (FER) over the 13 assay plates (A). Z-scores of FER over the 13 assay plates (B). Kernel density plot of Z-scores of FER with tertile cut-points for categorical analysis (C).

**A**

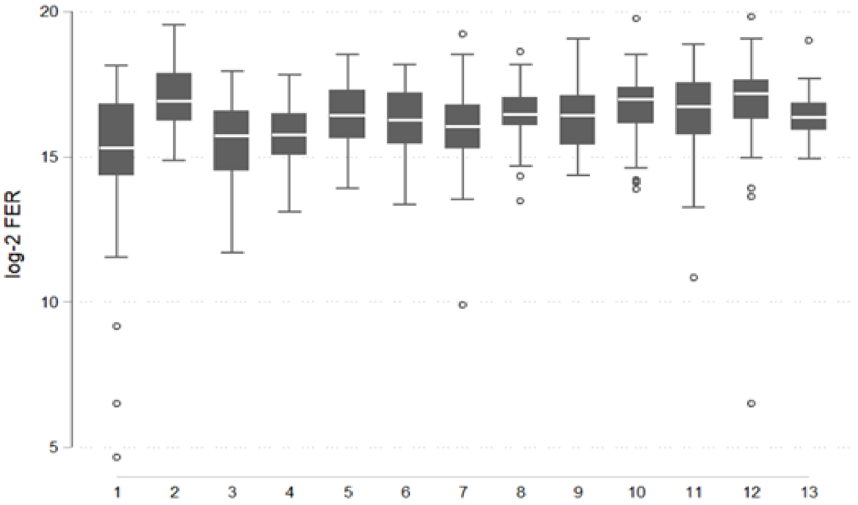

**B**

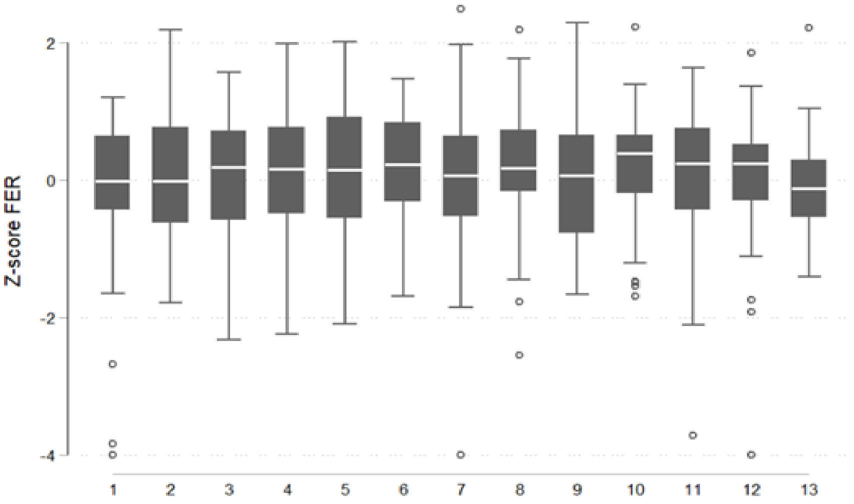

**C**

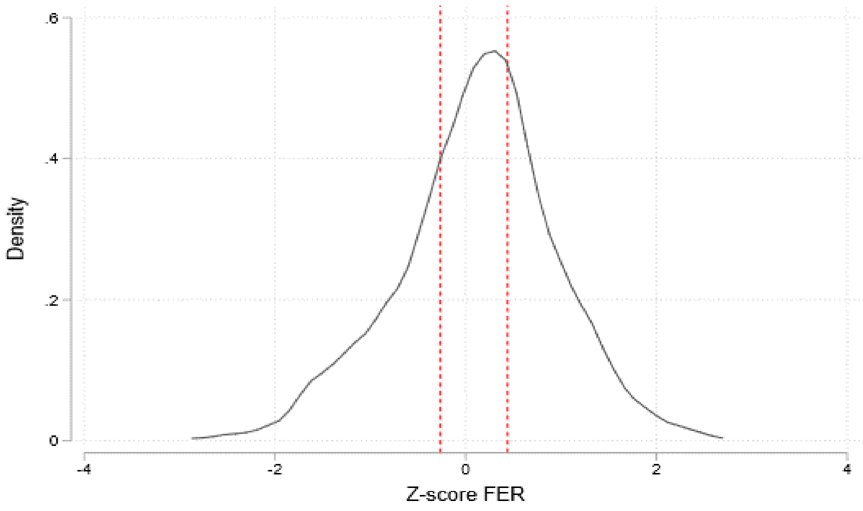

**Supplementary Figure 3.** The Spearman rho correlations of eight APP with each other, as measured in maternal serum samples of 429 unaffected controls. Abbreviations **A2M**: α-2-macroglobulin; **CRP**: C-reactive protein; **FER**: ferritin; **FIB**: fibrinogen; **PCT**: procalcitonin; **SAA**: serum amyloid A; **SAP**: serum amyloid P; and **tPA**: tissue plasminogen activator.

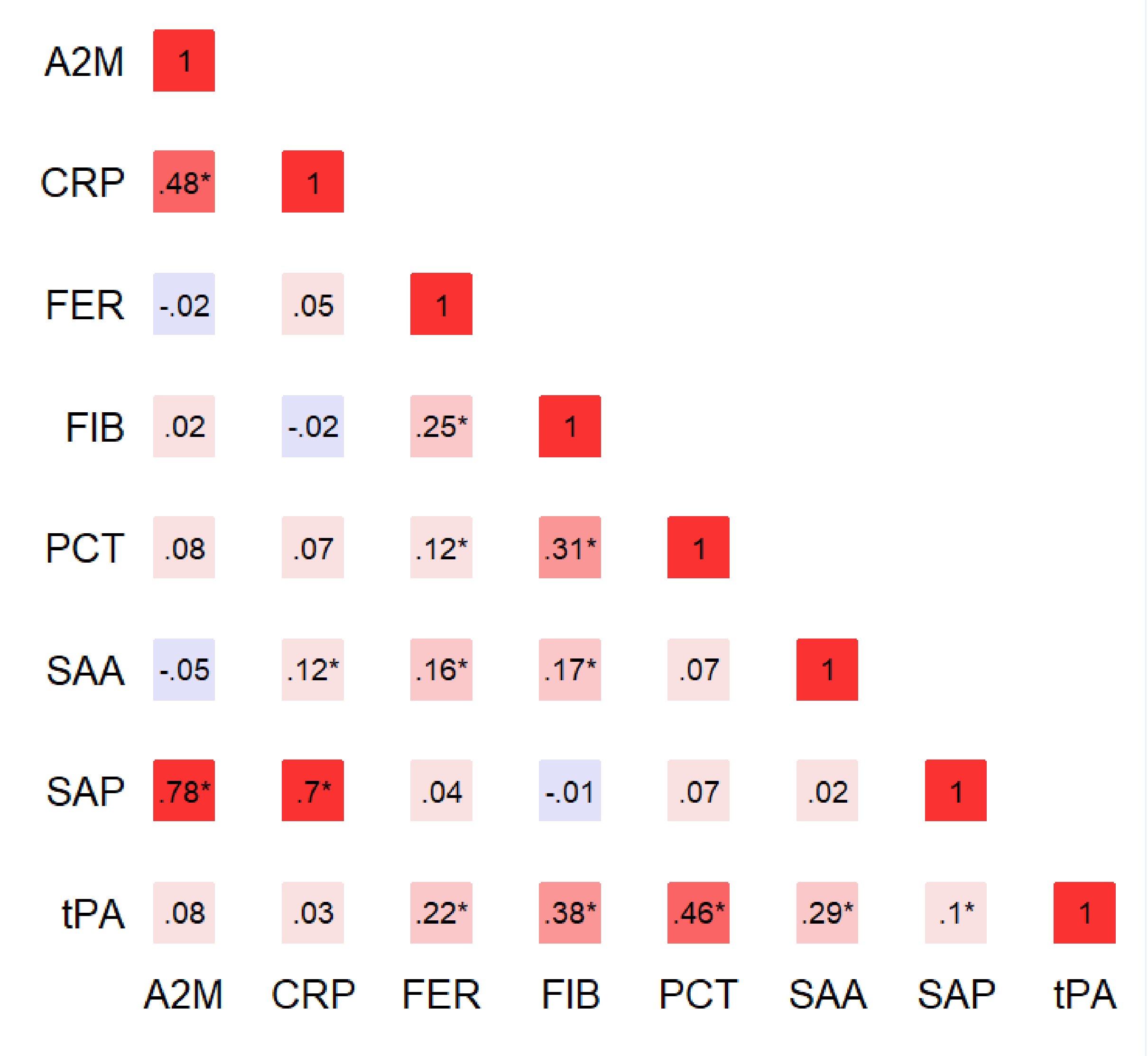

**Supplementary Figure 4.** Heat map showing the mean APP z-score by categories of the covariates, among 318 ASD-affected individuals in the cohort. Solid boxes indicate that the APP is associated with the covariate at p<0.05. Dashed boxes indicate that the APP is associated with the covariate at p<0.20. Abbreviations: **A2M**: α-2 macroglobulin; **CRP**: C-reactive protein; **FER**: ferritin; **FIB**: fibrinogen; **PCT**: procalcitonin; **SAA**: serum amyloid A; **SAP**: serum amyloid P; **tPA**: tissue plasminogen activator; Psych: Psychiatric; BMI: body mass index; Income Q: income quintile.

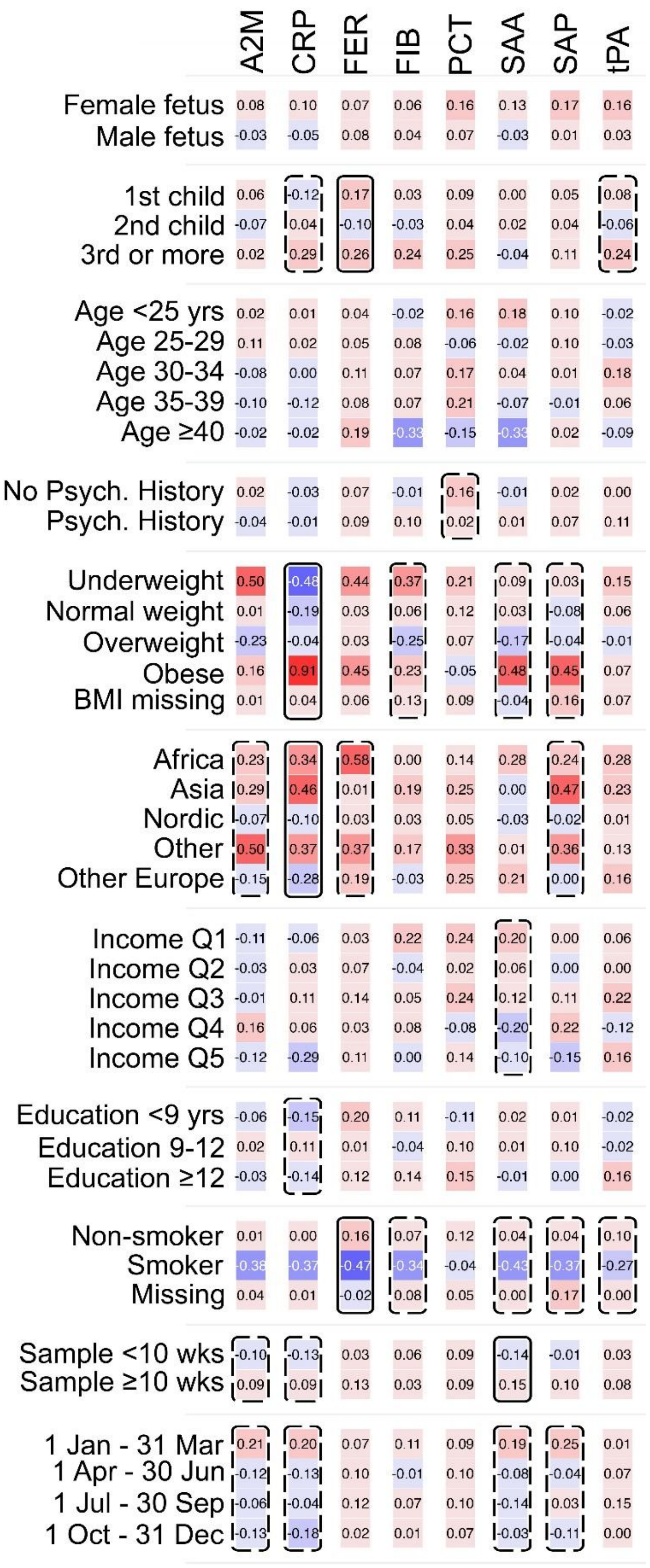

**Supplementary Figure 5**. Distribution of each APP measured in maternal serum samples, by ASD-case status.

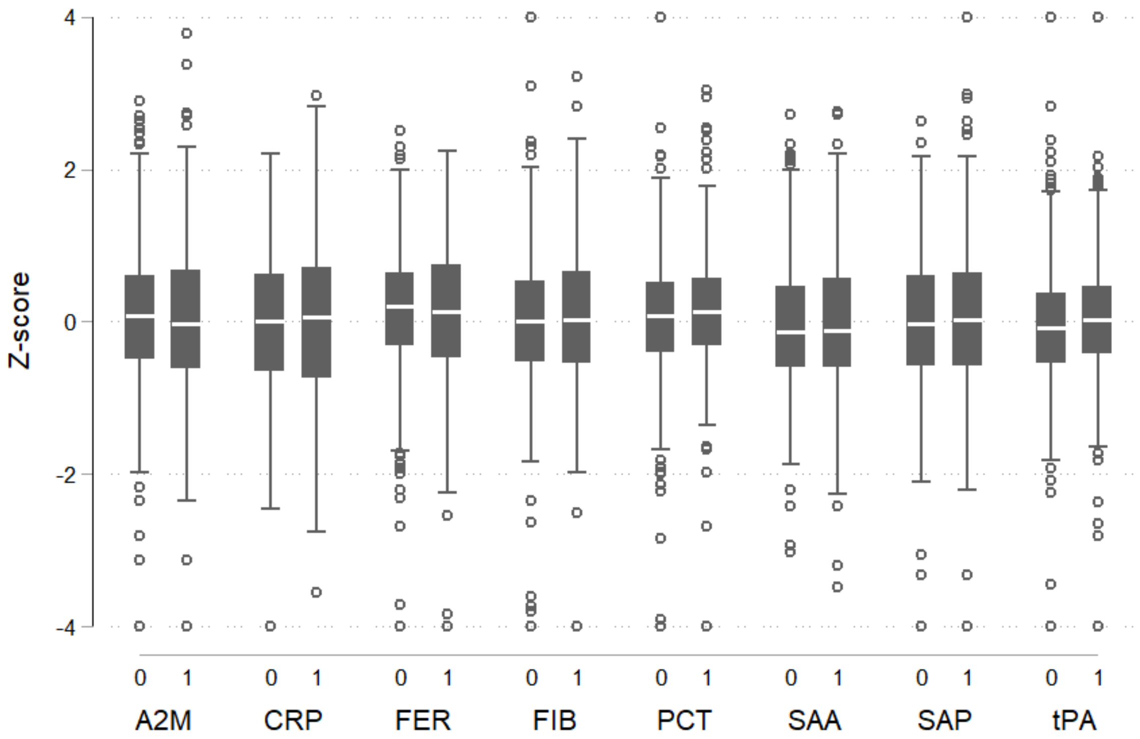

**Supplementary Figure 6**. The unadjusted relationship between APP and odds of ASD, stratified by co-occurrence of ID and ADHD, when comparing 318 ASD cases to 429 unaffected individuals selected from the cohort. Tertiles of each APP were created using the distribution of z-scores among unaffected individuals to set the cut-offs and the middle tertile was used as the referent category. P-values are shown for a Wald test with a null hypothesis that all APP categorical terms were jointly equal to zero, as a test of whether each APP was generally associated with the outcome. Abbreviations: **A2M**: α-2 macroglobulin; **CRP**: C-reactive protein; **FER**: ferritin; **FIB**: fibrinogen; **PCT**: procalcitonin; **SAA**: serum amyloid A; **SAP**: serum amyloid P; and **tPA**: tissue plasminogen activator.

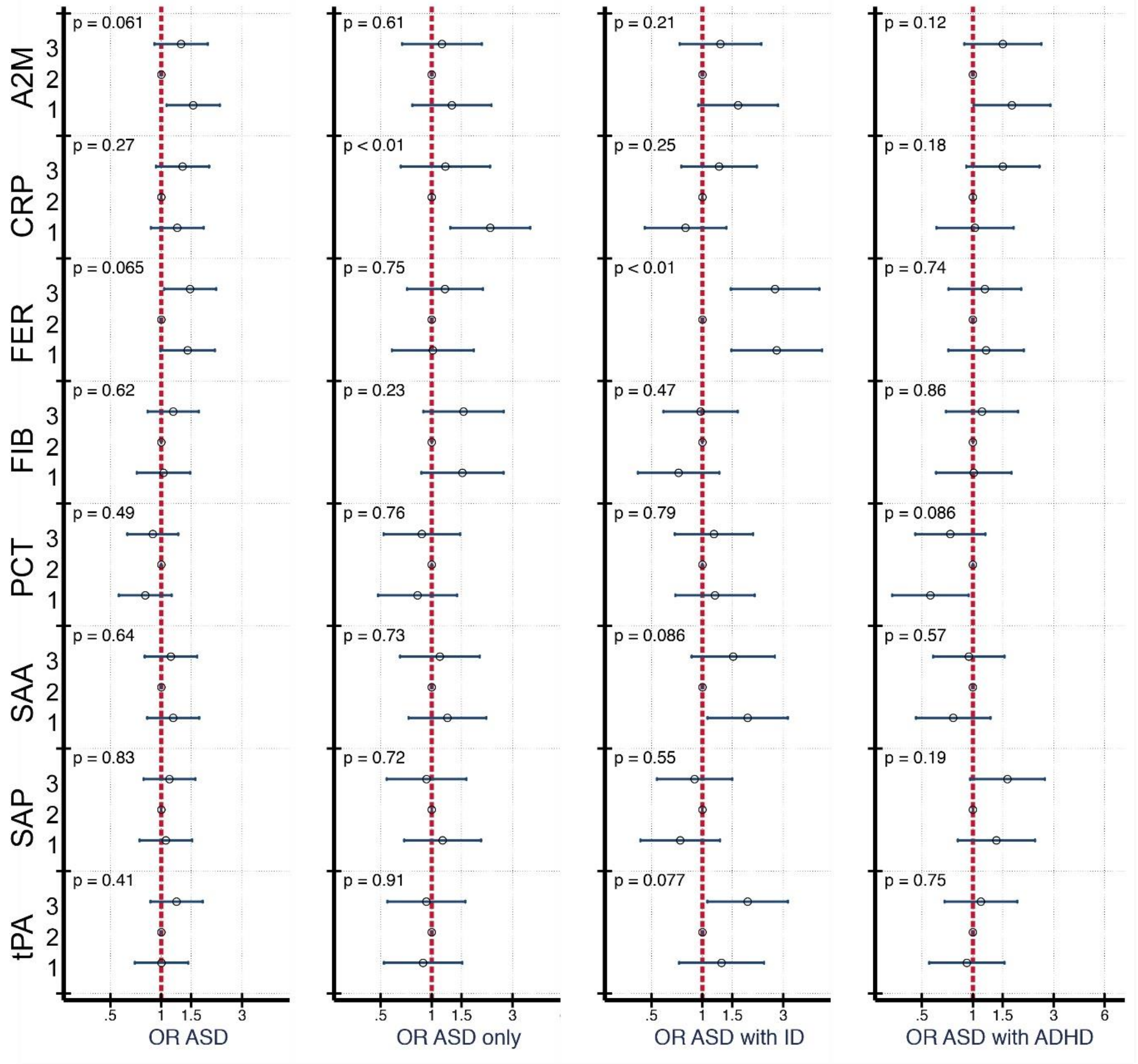

**Supplementary Figure 7.** The relationship between APP and odds of ASD with ID when comparing 101 individuals affected by ASD with co-occurring ID to 429 unaffected individuals selected from the cohort. Each panel displays the odds of ASD according to APP z-score, flexibly fit using restricted cubic spline models with three knots and a z-score=0 as the referent. The dashed line represents the unadjusted estimate of the relationship between each APP and odds of ASD. The solid line represents the fully adjusted model, adjusted for sex, birth order, maternal BMI, maternal psychiatric history, maternal region of origin, maternal age and family income quintile. The gray bands represent the 95% confidence interval for the fully adjusted model. P-values are shown for a Wald test with a null hypothesis that all APP spline terms were jointly equal to zero, as a test of whether each APP was generally associated with the outcome. Abbreviations: **A2M**: α-2 macroglobulin; **CRP**: C-reactive protein; **FER**: ferritin; **FIB**: fibrinogen; **PCT**: procalcitonin; **SAA**: serum amyloid A; **SAP**: serum amyloid P; and **tPA**: tissue plasminogen activator.

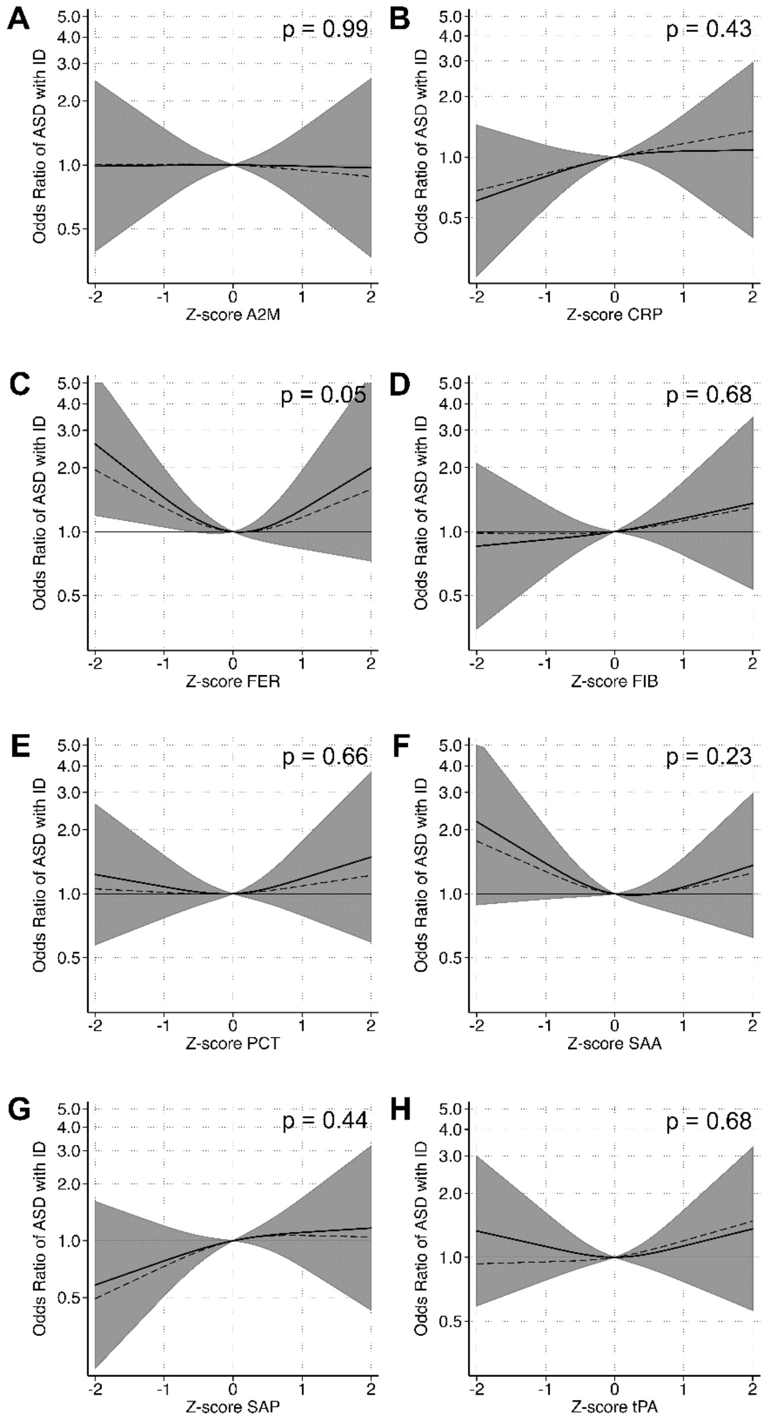

**Supplementary Figure 8**. The relationship between APP and odds of ASD without co-occurring ID or ADHD when comparing 100 individuals affected by ASD without co-occurring ID or ADHD to 429 unaffected individuals selected from the cohort. Each panel displays the odds of ASD according to APP z-score, flexibly fit using restricted cubic spline models with three knots and a z-score=0 as the referent. The dashed line represents the unadjusted estimate of the relationship between each APP and odds of ASD. The solid line represents the fully adjusted model, adjusted for sex, birth order, maternal BMI, maternal psychiatric history, maternal region of origin, maternal age and family income quintile. The gray bands represent the 95% confidence interval for the fully adjusted model. P-values are shown for a Wald test with a null hypothesis that all APP spline terms were jointly equal to zero, as a test of whether each APP was generally associated with the outcome. Abbreviations: **A2M**: α-2 macroglobulin; **CRP**: C-reactive protein; **FER**: ferritin; **FIB**: fibrinogen; **PCT**: procalcitonin; **SAA**: serum amyloid A; **SAP**: serum amyloid P; and **tPA**: tissue plasminogen activator.

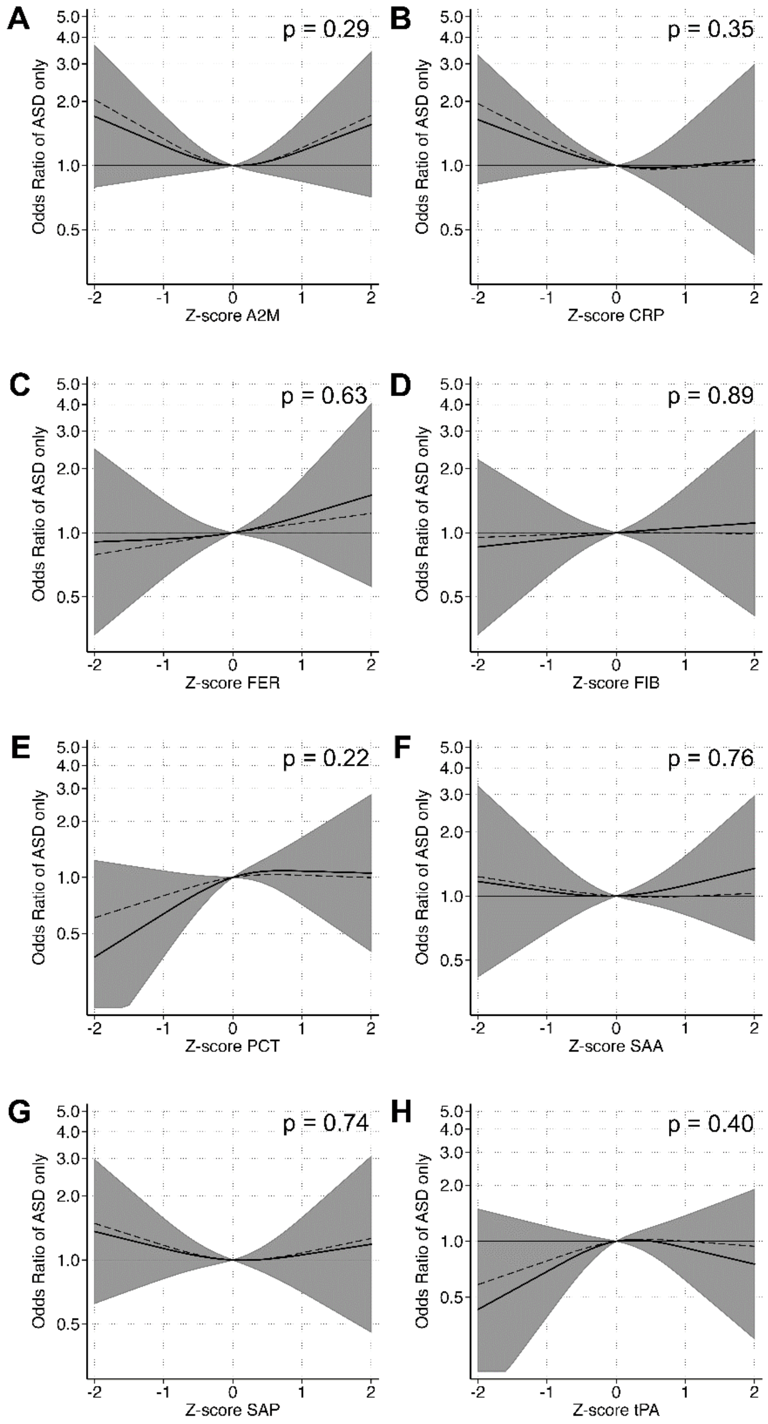

**Supplementary Figure 9**. The relationship between APP and odds of ASD with co-occurring ADHD when comparing 117 individuals affected by ASD with co-occurring ADHD to 429 unaffected individuals selected from the cohort. Each panel displays the odds of ASD according to APP z-score, flexibly fit using restricted cubic spline models with three knots and a z-score=0 as the referent. The dashed line represents the unadjusted estimate of the relationship between each APP and odds of ASD. The solid line represents the fully adjusted model, adjusted for sex, birth order, maternal BMI, maternal psychiatric history, maternal region of origin, maternal age and family income quintile. The gray bands represent the 95% confidence interval for the fully adjusted model. P-values are shown for a Wald test with a null hypothesis that all APP spline terms were jointly equal to zero, as a test of whether each APP was generally associated with the outcome. Abbreviations: **A2M**: α-2 macroglobulin; **CRP**: C-reactive protein; **FER**: ferritin; **FIB**: fibrinogen; **PCT**: procalcitonin; **SAA**: serum amyloid A; **SAP**: serum amyloid P; and **tPA**: tissue plasminogen activator.

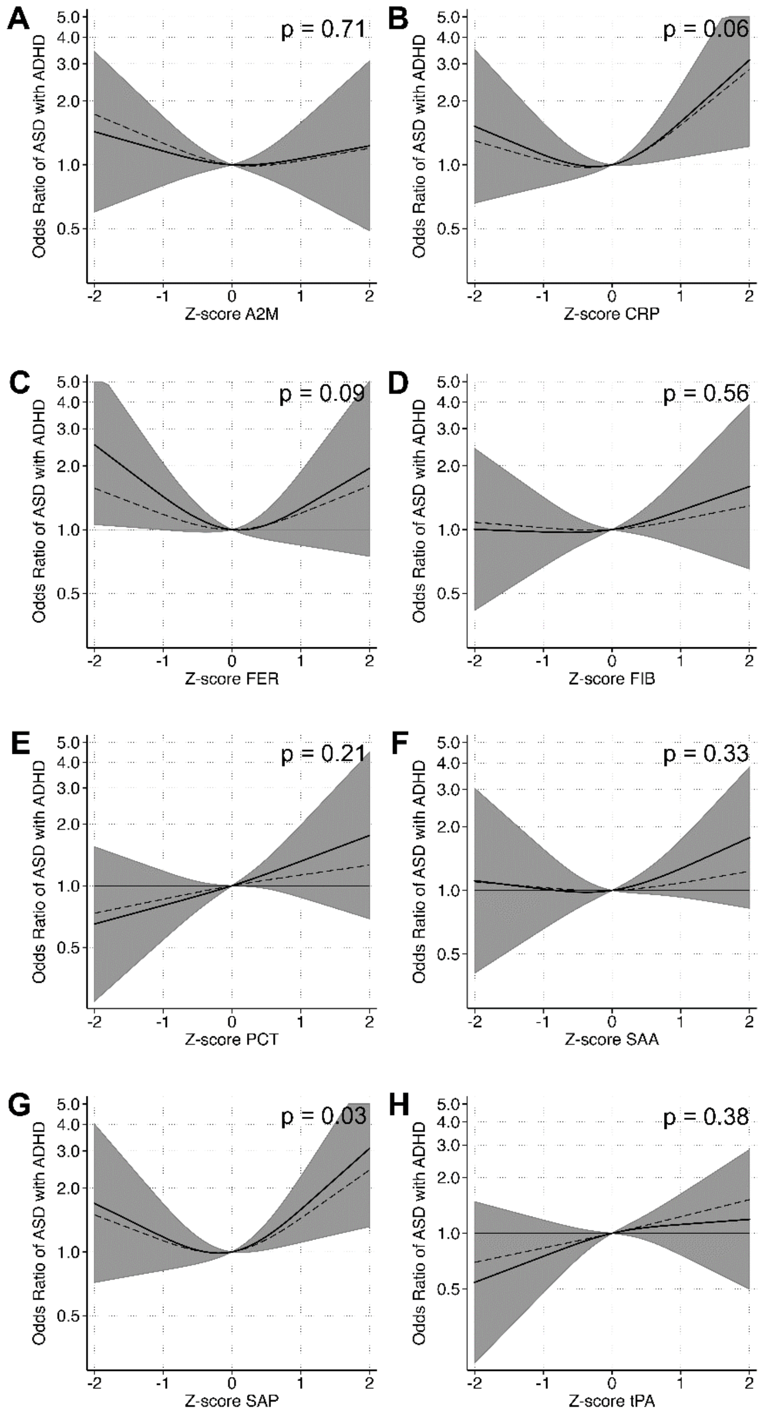

**Supplementary Figure 10.** The relationship between APP and odds of ASD, stratified by co-occurrence of ID and ADHD, when comparing 250 ASD cases to 349 unaffected individuals selected from the cohort. As a sensitivity analysis, the cohort was restricted to Nordic-born mothers. Tertiles of each APP were created using the distribution of z-scores among unaffected individuals to set the cut-offs and the middle tertile was used as the referent category. Models were adjusted for sex, birth order, maternal BMI, maternal psychiatric history, maternal region of origin, maternal age and family income quintile. P-values are shown for a Wald test with a null hypothesis that all APP categorical terms were jointly equal to zero, as a test of whether each APP was generally associated with the outcome. Abbreviations: **A2M**: α-2 macroglobulin; **CRP**: C-reactive protein; **FER**: ferritin; **FIB**: fibrinogen; **PCT**: procalcitonin; **SAA**: serum amyloid A; **SAP**: serum amyloid P; and **tPA**: tissue plasminogen activator.

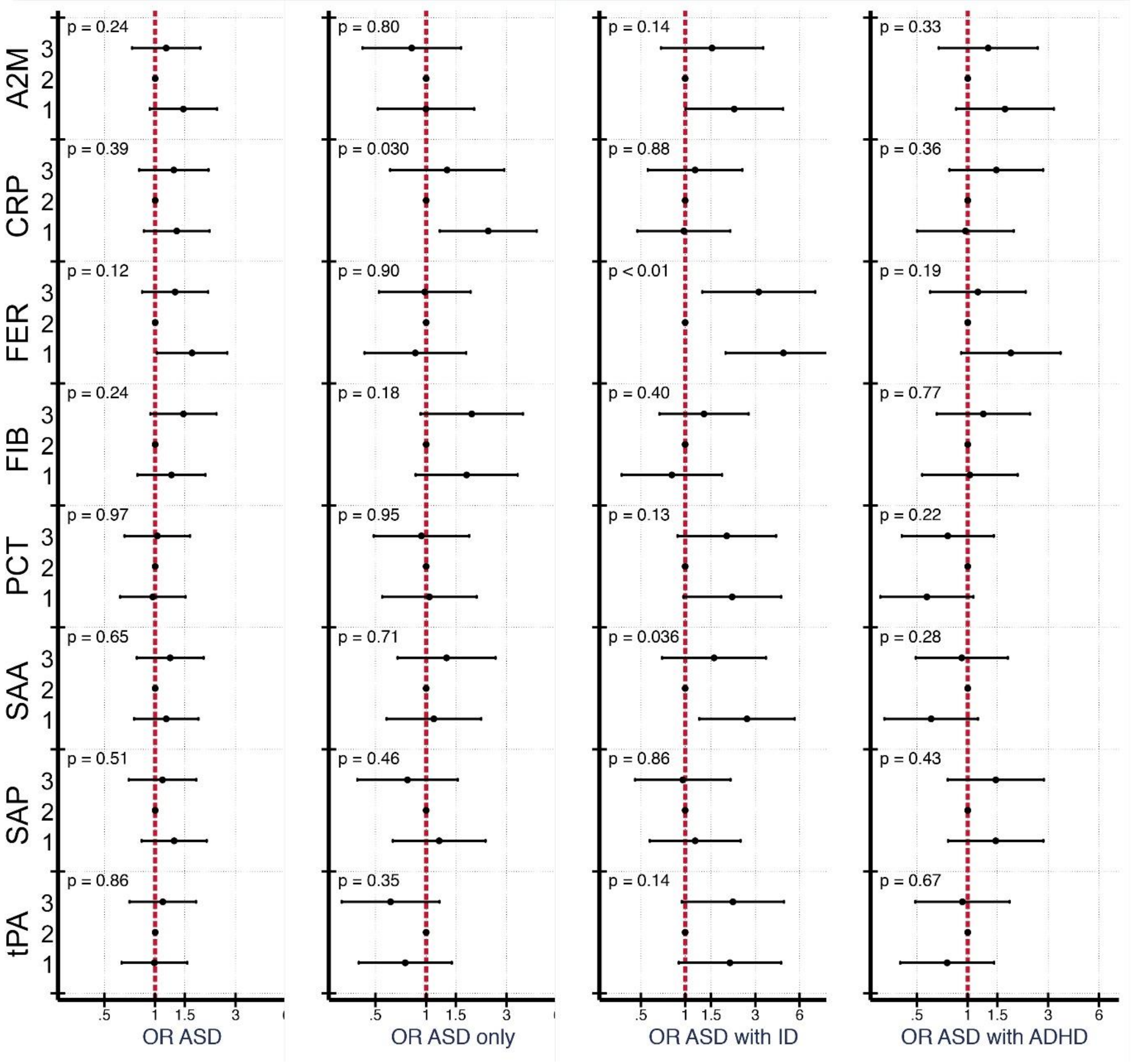

**Supplementary Figure 11.** The relationship between APP and odds of ASD, stratified by co-occurrence of ID and ADHD, when comparing 250 ASD cases to 349 unaffected individuals selected from the cohort. As a sensitivity analysis, regression models were adjusted for annual quarter at serum sample and gestational week at serum sample, in addition to covariates in the main analysis. Tertiles of each APP were created using the distribution of z-scores among unaffected individuals to set the cut-offs and the middle quintile was used as the referent category. P-values are shown for a Wald test with a null hypothesis that all APP categorical terms were jointly equal to zero, as a test of whether each APP was generally associated with the outcome. Abbreviations: **A2M**: α-2 macroglobulin; **CRP**: C-reactive protein; **FER**: ferritin; **FIB**: fibrinogen; **PCT**: procalcitonin; **SAA**: serum amyloid A; **SAP**: serum amyloid P; and **tPA**: tissue plasminogen activator.

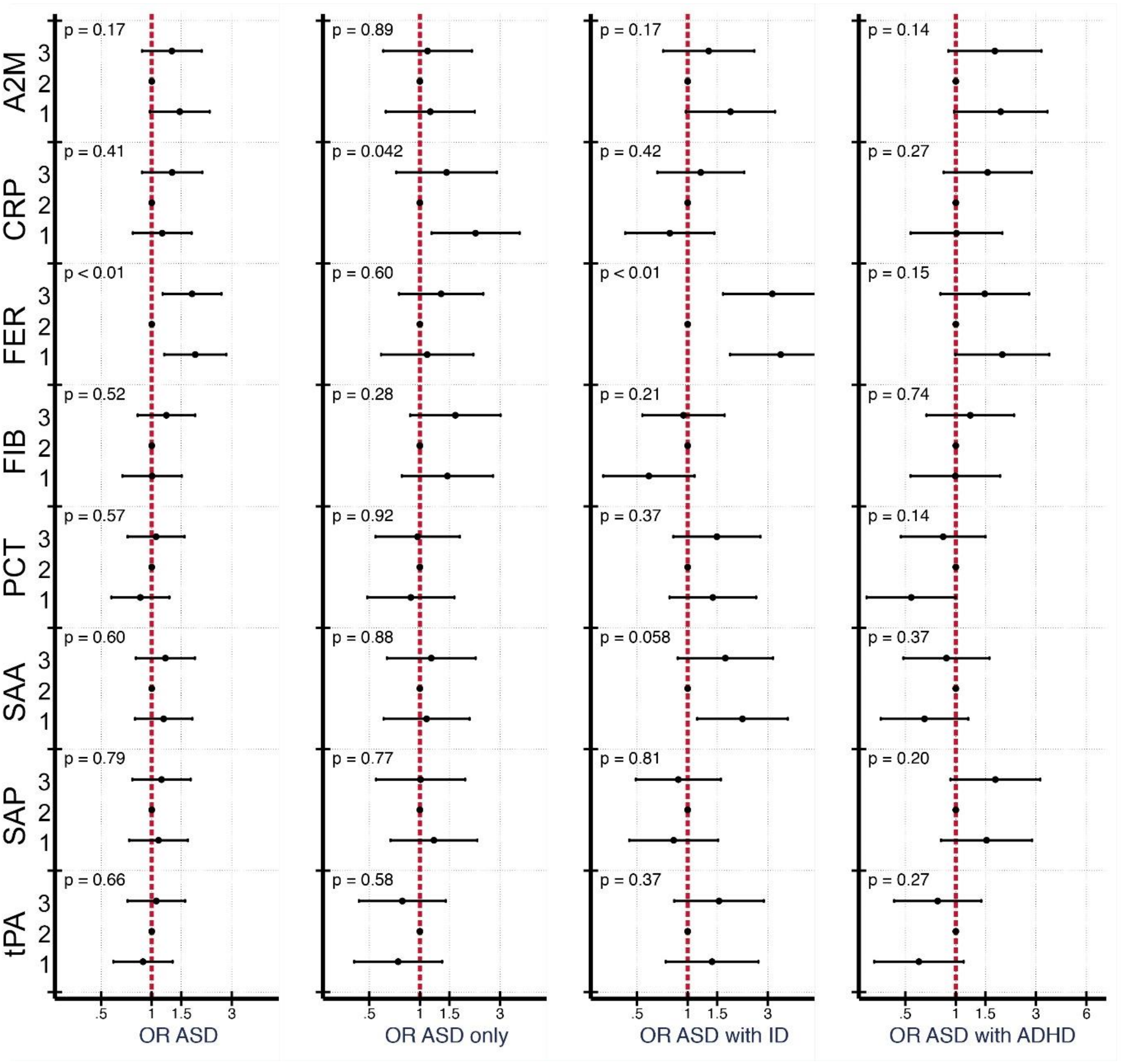

**Supplementary Table 1.** Characteristics of individuals in the source population (SYC Source Population), those for whom neonatal dried blood spots were selected (NDBS Cohort), those in NDBS cohort who were selected for APP-analysis (NDBS APP Cohort), those for whom maternal serum samples were selected (MS Cohort), and those for whom maternal serum samples were selected for APP analysis (MS Analysis Cohort).

| | ***SYC Source Cohort*[1]** | ***NDBS Cohort*[2]** | ***NDBS APP Cohort*[3]** | ***MS Cohort*[4]** | ***MS Analysis Cohort*[5]** |
|---|---|---|---|---|---|
| | **N = 98 597** | **N = 1 847** | **N = 1 154** | **N=559** | **N=438** |
| **Sex** | | | | | |
| Female | 48 158 (48.8%) | 879 (47.6%) | 556 (48.2%) | 259 (46.3%) | 206 (47.0%) |
| Male | 50 439 (51.2%) | 968 (52.4%) | 598 (51.8%) | 300 (53.7%) | 232 (53.0%) |
| **Birth Order** | | | | | |
| 1st born | 40 501 (41.1%) | 770 (41.7%) | 478 (41.4%) | 229 (41.0%) | 183 (41.8%) |
| 2nd born | 34 238 (34.7%) | 648 (35.1%) | 401 (34.7%) | 202 (36.1%) | 164 (37.4%) |
| 3rd or higher | 17 511 (17.8%) | 347 (18.8%) | 213 (18.5%) | 98 (17.5%) | 74 (16.9%) |
| missing | 6347 (6.4%) | 82 (4.4%) | 62 (5.4%) | 30 (5.4%) | 17 (3.9%) |
| **Maternal Age (years)** | | | | | |
| >25 | 13 767 (14.0%) | 251 (13.6%) | 156 (13.5%) | 66 (11.8%) | 45 (10.3%) |
| 25-29 | 29 684 (30.1%) | 572 (31.0%) | 322 (27.9%) | 149 (26.7%) | 112 (25.6%) |
| 30-34 | 35 526 (36.0%) | 634 (34.3%) | 406 (35.2%) | 218 (39.0%) | 179 (40.9%) |
| 35-39 | 16 302 (16.5%) | 336 (18.2%) | 235 (20.4%) | 112 (20.0%) | 90 (20.5%) |
| ≥40 | 3 161 (3.2%) | 54 (2.9%) | 35 (3.0%) | 14 (2.5%) | 12 (2.7%) |
| missing | 157 (0.2%) | | | | |
| **Maternal Psychiatric History** | | | | | |
| No | 65 155 (66.1%) | 1 210 (65.5%) | 756 (65.5%) | 380 (68.0%) | 292 (66.7%) |
| Yes | 33 288 (33.8%) | 637 (34.5%) | 398 (34.5%) | 179 (32.0%) | 146 (33.3%) |
| missing | 154 (0.2%) | | | | |
| **Maternal BMI (kg/m$^2$)** | | | | | |
| Normal | 49 181 (49.9%) | 935 (50.6%) | 564 (48.9%) | 265 (47.4%) | 216 (49.3%) |
| Underweight | 2252 (2.3%) | 39 (2.1%) | 23 (2.0%) | 9 (1.6%) | 8 (1.8%) |
| Overweight | 15 084 (15.3%) | 308 (16.7%) | 197 (17.1%) | 85 (15.2%) | 63 (14.4%) |
| Obese | 4 780 (4.8%) | 99 (5.4%) | 52 (4.5%) | 24 (4.3%) | 16 (3.7%) |

| | | | | | |
|---|---|---|---|---|---|
| missing | 27 300 (27.7%) | 466 (25.2%) | 318 (27.6%) | 176 (31.5%) | 135 (30.8%) |
| **Maternal Region of Birth** | | | | | |
| Africa | 4 807 (4.9%) | 103 (5.6%) | 67 (5.8%) | 33 (5.9%) | 18 (4.1%) |
| Asia | 10 960 (11.1%) | 186 (10.1%) | 114 (9.9%) | 55 (9.8%) | 34 (7.8%) |
| Nordic | 76 086 (77.2%) | 1 430 (77.4%) | 888 (76.9%) | 430 (76.9%) | 357 (81.5%) |
| Other | 2 523 (2.6%) | 52 (2.8%) | 38 (3.3%) | 20 (3.6%) | 16 (3.7%) |
| Other Europe | 4 067 (4.1%) | 76 (4.1%) | 47 (4.1%) | 21 (3.8%) | 13 (3.0%) |
| missing | 154 (0.16%) | | | | |
| **Family Income Quintile** | | | | | |
| 1 | 14 221 (14.4%) | 267 (14.5%) | 161 (14.0%) | 67 (12.0%) | 42 (9.6%) |
| 2 | 20 209 (20.5%) | 388 (21.0%) | 248 (21.5%) | 115 (20.6%) | 79 (18.0%) |
| 3 | 21 187 (21.5%) | 398 (21.5%) | 253 (21.9%) | 108 (19.3%) | 92 (21.0%) |
| 4 | 21 518 (21.8%) | 390 (21.1%) | 249 (21.6%) | 124 (22.2%) | 100 (22.8%) |
| 5 | 21 444 (21.7%) | 391 (21.2%) | 243 (21.1%) | 145 (25.9%) | 125 (28.5%) |
| missing | 18 (<1%) | 13 (0.7%) | | | |
| **Maternal Education Level at Birth** | | | | | |
| <9 years | 15 302 (15.5%) | 280 (15.2%) | 175 (15.2%) | 84 (15.0%) | 56 (12.8%) |
| 9-12 years | 44 067 (44.7%) | 820 (44.4%) | 501 (43.4%) | 230 (41.1%) | 186 (42.5%) |
| >12 | 38 647 (39.2%) | 727 (39.4%) | 475 (41.2%) | 242 (43.3%) | 195 (44.5%) |
| missing | 581 (0.6%) | 20 (1.1%) | 3 (0.3%) | 3 (0.5%) | 1 (0.2%) |
| **Trimester at First Antenatal Visit** | | | | | |
| 1 | 64 220 (65.1%) | 1 351 (73.1%) | 856 (74.2%) | 466 (83.4%) | 438 (100.0%) |
| 2 | 11 862 (12.0%) | 256 (13.9%) | 173 (15.0%) | 82 (14.7%) | |
| 3 | 1 246 (1.3%) | 22 (1.2%) | 15 (1.3%) | 10 (1.8%) | |
| missing | 21 269 (21.6%) | 218 (11.8%) | 110 (9.5%) | 1 (0.2%) | |
| **Number of Antenatal Visits** | | | | | |
| <9 | 14 362 (14.6%) | 270 (14.6%) | 189 (16.4%) | 95 (17.0%) | 54 (12.3%) |
| 9-13 | 47 819 (48.5%) | 909 (49.2%) | 589 (51.0%) | 278 (49.7%) | 238 (54.3%) |

| | | | | | |
|---|---|---|---|---|---|
| ≥14 | 8 613 (8.7%) | 158 (8.6%) | 81 (7.0%) | 30 (5.4%) | 28 (6.4%) |
| | 27 803 (28.2%) | 510 (27.6%) | 295 (25.6%) | 156 (27.9%) | 118 6.9% ) |

1. Cohort selected from the Stockholm Youth Cohort (SYC), from those born in Sweden between 1996 and 2000, and resident in Stockholm County for at least four years. See Supplementary Figure 1 for corresponding flowchart of sample selection.
2. Individuals selected for collection of neonatal dried blood spot (NDBS) sample as a random sample from the larger SYC cohort. See Supplementary Figure 1.
3. Individuals selected for analysis of acute phase proteins in NDBS samples. See Supplementary Figure 1.
4. Individuals for whom maternal serum samples were collected. See Supplementary Figure 1.
5. Individuals for whom maternal serum samples were selected for analysis of acute phase proteins. See Supplementary Figure 1.

**Supplementary Table 2.** Quality control statistics for multiplex assays to analyze acute phase protein concentrations in maternal serum samples.

| Analyte | %CV (controls)[1] | Below LLOQ | LLOQ[2] | Above ULOQ | ULOQ[3] | Imputed[4] |
|---|---|---|---|---|---|---|
| α-2-macroglobulin | 15.8 | 3 (0.40%) | 0.61 ng/ml | 3 (0.40%) | 4062.09 ng/ml | 6 (0.80%) |
| C-reactive protein | 13.9 | 3 (0.40%) | 0.01 ng/ml | 1 (0.13%) | 57 ng/ml | 4 (0.54%) |
| Ferritin | 20.5 | 3 (0.40%) | 2.49 pg/ml | 0 (0.00%) | 39405.2 pg/ml | 3 (0.40%) |
| Fibrinogen | 11.2 | 4 (0.54%) | 3.01 ng/ml | 0 (0.00%) | 758.4 ng/ml | 4 (0.54%) |
| Haptoglobin | 16.2 | 2 (0.27%) | 0.04 ng/ml | 564 (75.50%) | 277.86 ng/ml | 566 (75.77%) |
| Procalcitonin | 34.5 | 5 (0.67%) | 4.4 pg/ml | 0 (0.00%) | 5365.04 pg/ml | 5 (0.67%) |
| Serum amyloid A | 18.7 | 5 (0.67%) | 0.29 ng/ml | 118 (15.80%) | 388.44 ng/ml | 123 (16.47%) |
| Serum amyloid P | 11.4 | 3 (0.40%) | 0.01 ng/ml | 36 (4.82%) | 186.54 ng/ml | 39 (5.22%) |
| Tissue plasminogen activator | 25.5 | 4 (0.54%) | 1.95 pg/ml | 0 (0.00%) | 4487.54 pg/ml | 4 (0.54%) |
| Average | | 0.48% | | 10.74% | | 11.21% |

1. The percent CV is based on standardized controls that were run in duplicate on each of the 13 assay plates.
2. Lower Limit of Quantitation, average across 13 assay plates.
3. Upper Limit of Quantitation, average across 13 assay plates.
4. The absolute and relative frequency of observations that were imputed, including those values that were below the LLOQ, above the ULOQ, or that the Bio-plex Manager software indicated were near the LLOQ and thus less certain than values higher in the range of quantitation.

**Supplementary Table 3**. P-values for the association of maternal APP with other covariates among 429 unaffected individuals. We examined the association of each covariate with the APP z-scores by regressing each APP over the categories of the covariates, followed by a joint Wald test of the hypothesis that all coefficients for the categorical indicators are equal to zero, a test whether each APP was generally associated with the covariate.

| | *A2M* | *CRP* | *FER* | *FIB* | *PCT* | *SAA* | *SAP* | *tPA* |
|---|---|---|---|---|---|---|---|---|
| *Sex* | 0.169 | 0.287 | 0.169 | 0.323 | 0.724 | 0.705 | 0.690 | 0.396 |
| *Birth Order* | **0.044** | 0.145 | 0.561 | 0.587 | 0.410 | 0.660 | 0.156 | 0.339 |
| *Maternal Age* | 0.250 | 0.165 | 0.329 | 0.492 | 0.256 | 0.146 | 0.307 | 0.212 |
| *Maternal Psych. Hist.* | 0.525 | 0.669 | 0.330 | 0.311 | 0.637 | **0.029** | 0.983 | 0.190 |
| *Maternal BMI* | **0.049** | **<0.001** | 0.840 | 0.583 | 0.320 | 0.259 | **0.039** | 0.540 |
| *Maternal Country of Birth* | 0.872 | 0.285 | 0.068 | 0.273 | 0.092 | 0.557 | 0.752 | 0.436 |
| *Parental Income* | 0.740 | 0.131 | 0.225 | 0.532 | 0.088 | 0.399 | 0.571 | 0.054 |
| *Maternal Education at Birth* | 0.512 | 0.087 | 0.434 | 0.142 | 0.061 | 0.801 | 0.604 | 0.986 |
| *Maternal Smoking* | 0.262 | 0.755 | 0.112 | 0.220 | 0.198 | 0.151 | 0.439 | 0.547 |
| *Gest. Age at Sampling* | 0.276 | **0.001** | 0.852 | 0.502 | 0.896 | 0.660 | 0.189 | 0.975 |
| *Sampling Quarter* | 0.817 | 0.745 | 0.473 | 0.551 | 0.989 | 0.916 | 0.996 | 0.224 |

Abbreviations: **Psych**: Psychiatric; **Hist**; History; **BMI**: Body Mass Index; **A2M**: α-2 macroglobulin; **CRP**: C-reactive protein; **FER**: ferritin; **FIB**: fibrinogen; **PCT**: procalcitonin; **SAA**: serum amyloid A; **SAP**: serum amyloid P; and **tPA**: tissue plasminogen activator.

**Supplementary Table 4**. P-values for the association of maternal APP with other covariates among 318 ASD affected individuals. We examined the association of each covariate with the APP z-scores by regressing each APP over the categories of the covariates, followed by a joint Wald test of the hypothesis that all coefficients for the categorical indicators are equal to zero, a test whether each APP was generally associated with the covariate.

| | *A2M* | *CRP* | *FER* | *FIB* | *PCT* | *SAA* | *SAP* | *tPA* |
|---|---|---|---|---|---|---|---|---|
| *Sex* | 0.393 | 0.308 | 0.954 | 0.897 | 0.502 | 0.253 | 0.265 | 0.302 |
| *Birth Order* | 0.575 | 0.074 | **0.044** | 0.264 | 0.435 | 0.954 | 0.933 | 0.167 |
| *Maternal Age* | 0.656 | 0.942 | 0.973 | 0.579 | 0.228 | 0.528 | 0.941 | 0.497 |
| *Maternal Psych. Hist.* | 0.563 | 0.832 | 0.833 | 0.276 | 0.177 | 0.848 | 0.622 | 0.334 |
| *Maternal BMI* | 0.280 | **<0.001** | 0.340 | 0.084 | 0.944 | 0.161 | 0.116 | 0.984 |
| *Maternal Country of Birth* | 0.062 | **0.016** | 0.102 | 0.800 | 0.554 | 0.724 | **0.047** | 0.479 |
| *Parental Income* | 0.525 | 0.263 | 0.967 | 0.688 | 0.205 | 0.197 | 0.273 | 0.208 |
| *Maternal Education at Birth* | 0.512 | 0.087 | 0.434 | 0.142 | 0.061 | 0.801 | 0.604 | 0.986 |
| *Maternal Smoking* | 0.235 | 0.317 | **0.011** | 0.147 | 0.705 | 0.143 | 0.094 | 0.196 |
| *Gest. Age at Sampling* | 0.094 | 0.067 | 0.356 | 0.722 | 0.998 | **0.011** | 0.336 | 0.611 |
| *Sampling Quarter* | 0.078 | 0.094 | 0.933 | 0.829 | 0.997 | 0.162 | 0.097 | 0.770 |

Abbreviations: **Psych**: Psychiatric; **Hist**; History; **BMI**: Body Mass Index; **A2M**: α-2 macroglobulin; **CRP**: C-reactive protein; **FER**: ferritin; **FIB**: fibrinogen; **PCT**: procalcitonin; **SAA**: serum amyloid A; **SAP**: serum amyloid P; and **tPA**: tissue plasminogen activator.

**Supplementary Table 5.** P-values for non-linearity in restricted cubic spline analyses. Not all relationships we considered are necessarily non-linear. To assess evidence for
non-linear relationships, we tested the null-hypothesis that all spline terms that would indicate a change in the slope of the relationship (i.e., all but the first spline term) are equal to zero using a Wald test.

| | ***Any ASD*** | | ***ASD only*** | | ***ASD w/ ID*** | | ***ASD w/ ADHD*** | |
|---|---|---|---|---|---|---|---|---|
| | unadjusted | adjusted | unadjusted | adjusted | unadjusted | adjusted | unadjusted | adjusted |
| *A2M* | 0.099 | 0.263 | **0.028** | 0.120 | 0.867 | 0.956 | 0.233 | 0.454 |
| *CRP* | 0.083 | 0.200 | 0.291 | 0.445 | 0.905 | 0.597 | **0.034** | **0.038** |
| *FER* | 0.090 | **0.028** | 0.970 | 0.714 | 0.084 | **0.030** | 0.133 | **0.039** |
| *FIB* | 0.651 | 0.555 | 0.929 | 0.953 | 0.699 | 0.846 | 0.566 | 0.523 |
| *PCT* | 0.871 | 0.739 | 0.492 | 0.301 | 0.691 | 0.377 | 0.902 | 0.854 |
| *SAA* | 0.298 | 0.170 | 0.717 | 0.548 | 0.197 | 0.123 | 0.615 | 0.360 |
| *SAP* | 0.208 | 0.188 | 0.343 | 0.508 | 0.415 | 0.653 | **0.034** | **0.022** |
| *tPA* | 0.990 | 0.994 | 0.402 | 0.207 | 0.584 | 0.384 | 0.921 | 0.561 |

Abbreviations: **ASD**: autism spectrum disorders; **ADHD**: attention-deficit/hyperactivity disorder; **ID**: intellectual disability**; A2M**: α-2 macroglobulin; **CRP**: C-reactive protein; **FER**: ferritin; **FIB**: fibrinogen; **PCT**: procalcitonin; **SAA**: serum amyloid A; **SAP**: serum amyloid P; and **tPA**: tissue plasminogen activator.

STROBE Statement—Checklist of items that should be included in reports of ***case-control studies***

| | Item No | Recommendation | Page No |
|---|---|---|---|
| **Title and abstract** | 1 | (*a*) Indicate the study's design with a commonly used term in the title or the abstract | 1 |
| | | (*b*) Provide in the abstract an informative and balanced summary of what was done and what was found | 1 |
| **Introduction** | | | |
| Background/rationale | 2 | Explain the scientific background and rationale for the investigation being reported | 2 |
| Objectives | 3 | State specific objectives, including any prespecified hypotheses | 2 |
| **Methods** | | | |
| Study design | 4 | Present key elements of study design early in the paper | 3 |
| Setting | 5 | Describe the setting, locations, and relevant dates, including periods of recruitment, exposure, follow-up, and data collection | 3 |
| Participants | 6 | (*a*) Give the eligibility criteria, and the sources and methods of case ascertainment and control selection. Give the rationale for the choice of cases and controls | 3 |
| | | (*b*) For matched studies, give matching criteria and the number of controls per case | |
| Variables | 7 | Clearly define all outcomes, exposures, predictors, potential confounders, and effect modifiers. Give diagnostic criteria, if applicable | 3-5 |
| Data sources/ measurement | 8* | For each variable of interest, give sources of data and details of methods of assessment (measurement). Describe comparability of assessment methods if there is more than one group | 3-5 |
| Bias | 9 | Describe any efforts to address potential sources of bias | 4-5 |
| Study size | 10 | Explain how the study size was arrived at | 3 |
| Quantitative variables | 11 | Explain how quantitative variables were handled in the analyses. If applicable, describe which groupings were chosen and why | 4-5 |
| Statistical methods | 12 | (*a*) Describe all statistical methods, including those used to control for confounding | 4-5 |
| | | (*b*) Describe any methods used to examine subgroups and interactions | 5 |
| | | (*c*) Explain how missing data were addressed | 4 |
| | | (*d*) If applicable, explain how matching of cases and controls was addressed | |
| | | (*e*) Describe any sensitivity analyses | 5 |
| **Results** | | | |

| | | | |
|---|---|---|---|
| Participants | 13* | (a) Report numbers of individuals at each stage of study—eg numbers potentially eligible, examined for eligibility, confirmed eligible, included in the study, completing follow-up, and analysed | 3 |
| | | (b) Give reasons for non-participation at each stage | 3 |
| | | (c) Consider use of a flow diagram | 3 |
| Descriptive data | 14* | (a) Give characteristics of study participants (eg demographic, clinical, social) and information on exposures and potential confounders | 5 |
| | | (b) Indicate number of participants with missing data for each variable of interest | 4-5 |
| Outcome data | 15* | Report numbers in each exposure category, or summary measures of exposure | 5 |

| | | | |
|---|---|---|---|
| Main results | 16 | (*a*) Give unadjusted estimates and, if applicable, confounder-adjusted estimates and their precision (eg, 95% confidence interval). Make clear which confounders were adjusted for and why they were included | 6 |
| | | (*b*) Report category boundaries when continuous variables were categorized | 4,5 |
| | | (*c*) If relevant, consider translating estimates of relative risk into absolute risk for a meaningful time period | |
| Other analyses | 17 | Report other analyses done—eg analyses of subgroups and interactions, and sensitivity analyses | 6 |
| **Discussion** | | | |
| Key results | 18 | Summarise key results with reference to study objectives | 7 |
| Limitations | 19 | Discuss limitations of the study, taking into account sources of potential bias or imprecision. Discuss both direction and magnitude of any potential bias | 10-11 |
| Interpretation | 20 | Give a cautious overall interpretation of results considering objectives, limitations, multiplicity of analyses, results from similar studies, and other relevant evidence | 8-10 |
| Generalisability | 21 | Discuss the generalisability (external validity) of the study results | 10 |
| **Other information** | | | |
| Funding | 22 | Give the source of funding and the role of the funders for the present study and, if applicable, for the original study on which the present article is based | 11 |

*Give information separately for cases and controls.

**Note:** An Explanation and Elaboration article discusses each checklist item and gives methodological background and published examples of transparent reporting. The STROBE checklist is best used in conjunction with this article (freely available on the Web sites of PLoS Medicine at http://www.plosmedicine.org/, Annals of Internal Medicine at http://www.annals.org/, and Epidemiology at http://www.epidem.com/). Information on the STROBE Initiative is available at http://www.strobe-statement.org.

### ICMJE DISCLOSURE FORM

**Date: 2021-03-03_______________________________________________________________________**
**Your Name: Martin Brynge_____________________________________________________________________**
**Manuscript Title: Maternal Levels of Acute Phase Proteins in Early Pregnancy and Risk of Autism Spectrum Disorders in Offspring _____________________________________________________________________**
**Manuscript number (if known):_____________________________________________________________________**

**In the interest of transparency, we ask you to disclose all relationships/activities/interests listed below that are related to the content of your manuscript. "Related" means any relation with for-profit or not-for-profit third parties whose interests may be affected by the content of the manuscript. Disclosure represents a commitment to transparency and does not necessarily indicate a bias. If you are in doubt about whether to list a relationship/activity/interes it is preferable that you do so.**

**The following questions apply to the author's relationships/activities/interests as they relate to the current manuscript only.**

**The author's relationships/activities/interests should be defined broadly. For example, if your manuscript pertains to the epidemiology of hypertension, you should declare all relationships with manufacturers of antihypertensive medication, even if that medication is not mentioned in the manuscript.**

**In item #1 below, report all support for the work reported in this manuscript without time limit. For all other items, the time frame for disclosure is the past 36 months.**

| | | **Name all entities with whom you have this relationship or indicate none (add rows as needed)** | **Specifications/Comments (e.g., if payments were made to you or to your institution)** |
|---|---|---|---|
| | | **Time frame: Since the initial planning of the work** | |
| 1 | All support for the present manuscript (e.g., funding, provision of study materials, medical writing, article processing charges, etc.) **No time limit for this item.** | X____None | |
| | | **Time frame: past 36 months** | |
| 2 | Grants or contracts from any entity (if not indicated in item #1 above). | X____None | |
| 3 | Royalties or licenses | X____None | |
| 4 | Consulting fees | X____None | |
| 5 | Payment or honoraria for lectures, presentations, speakers bureaus, manuscript writing or educational events | X____None | |
| 6 | | X____None | |

| | | |
|---|---|---|
| | Payment for expert testimony | |
| 7 | Support for attending meetings and/or travel | X____None |
| 8 | Patents planned, issued or pending | X____None |
| 9 | Participation on a Data Safety Monitoring Board or Advisory Board | X____None |
| 10 | Leadership or fiduciary role in other board, society, committee or advocacy group, paid or unpaid | X____None |
| 11 | Stock or stock options | X____None |
| 12 | Receipt of equipment, materials, drugs, medical writing, gifts or other services | X____None |
| 13 | Other financial or non-financial interests | X____None |

**Please place an "X" next to the following statement to indicate your agreement:**

**X___ I certify that I have answered every question and have not altered the wording of any of the questions on this form.**

### ICMJE DISCLOSURE FORM

**Date: 2021-03-03______________________________________________________________________**
**Your Name: Renee Gardner________________________________________________________________**
**Manuscript Title: Maternal Levels of Acute Phase Proteins in Early Pregnancy and Risk of Autism Spectrum Disorders in Offspring _________________________________________________________________**
**Manuscript number (if known):_____________________________________________________________**

**In the interest of transparency, we ask you to disclose all relationships/activities/interests listed below that are related to the content of your manuscript. "Related" means any relation with for-profit or not-for-profit third parties whose interests may be affected by the content of the manuscript. Disclosure represents a commitment to transparency and does not necessarily indicate a bias. If you are in doubt about whether to list a relationship/activity/interes it is preferable that you do so.**

**The following questions apply to the author's relationships/activities/interests as they relate to the current manuscript only.**

**The author's relationships/activities/interests should be defined broadly. For example, if your manuscript pertains to the epidemiology of hypertension, you should declare all relationships with manufacturers of antihypertensive medication, even if that medication is not mentioned in the manuscript.**

**In item #1 below, report all support for the work reported in this manuscript without time limit. For all other items, the time frame for disclosure is the past 36 months.**

| | | **Name all entities with whom you have this relationship or indicate none (add rows as needed)** | **Specifications/Comments (e.g., if payments were made to you or to your institution)** |
|---|---|---|---|
| | | **Time frame: Since the initial planning of the work** | |
| 1 | All support for the present manuscript (e.g., funding, provision of study materials, medical writing, article processing charges, etc.) **No time limit for this item.** | ____None | |
| | | Swedish Research Council | |
| | | **Time frame: past 36 months** | |
| 2 | Grants or contracts from any entity (if not indicated in item #1 above). | X____None | |
| 3 | Royalties or licenses | X____None | |
| 4 | Consulting fees | X____None | |
| 5 | Payment or honoraria for lectures, presentations, speakers bureaus, manuscript writing or educational events | X____None | |
| 6 | | X____None | |

| | | |
|---|---|---|
| | Payment for expert testimony | |
| 7 | Support for attending meetings and/or travel | X____None |
| 8 | Patents planned, issued or pending | X____None |
| 9 | Participation on a Data Safety Monitoring Board or Advisory Board | X____None |
| 10 | Leadership or fiduciary role in other board, society, committee or advocacy group, paid or unpaid | X____None |
| 11 | Stock or stock options | X____None |
| 12 | Receipt of equipment, materials, drugs, medical writing, gifts or other services | X____None |
| 13 | Other financial or non-financial interests | X____None |

**Please place an "X" next to the following statement to indicate your agreement:**

**X___ I certify that I have answered every question and have not altered the wording of any of the questions on this form.**

**ICMJE DISCLOSURE FORM**

**Date: 2021-03-03________________________________________________________________________**
**Your Name: Håkan Karlsson___________________________________________________________________**
**Manuscript Title: Maternal Levels of Acute Phase Proteins in Early Pregnancy and Risk of Autism Spectrum Disorders in Offspring ______________________________________________________________________**
**Manuscript number (if known):_________________________________________________________________**

**In the interest of transparency, we ask you to disclose all relationships/activities/interests listed below that are related to the content of your manuscript. "Related" means any relation with for-profit or not-for-profit third parties whose interests may be affected by the content of the manuscript. Disclosure represents a commitment to transparency and does not necessarily indicate a bias. If you are in doubt about whether to list a relationship/activity/interes it is preferable that you do so.**

**The following questions apply to the author's relationships/activities/interests as they relate to the <u>current manuscript only</u>.**

**The author's relationships/activities/interests should be <u>defined broadly</u>. For example, if your manuscript pertains to the epidemiology of hypertension, you should declare all relationships with manufacturers of antihypertensive medication, even if that medication is not mentioned in the manuscript.**

**In item #1 below, report all support for the work reported in this manuscript without time limit. For all other items, the time frame for disclosure is the past 36 months.**

| | | **Name all entities with whom you have this relationship or indicate none (add rows as needed)** | **Specifications/Comments (e.g., if payments were made to you or to your institution)** |
|---|---|---|---|
| | | **Time frame: Since the initial planning of the work** | |
| 1 | All support for the present manuscript (e.g., funding, provision of study materials, medical writing, article processing charges, etc.) **No time limit for this item.** | ____None | |
| | | Stanley Medical Research Institute | |
| | | **Time frame: past 36 months** | |
| 2 | Grants or contracts from any entity (if not indicated in item #1 above). | X____None | |
| 3 | Royalties or licenses | X____None | |
| 4 | Consulting fees | X____None | |
| 5 | Payment or honoraria for lectures, presentations, speakers bureaus, manuscript writing or educational events | X____None | |

| | | |
|---|---|---|
| 6 | Payment for expert testimony | X____None |
| 7 | Support for attending meetings and/or travel | X____None |
| 8 | Patents planned, issued or pending | X____None |
| 9 | Participation on a Data Safety Monitoring Board or Advisory Board | X____None |
| 10 | Leadership or fiduciary role in other board, society, committee or advocacy group, paid or unpaid | X____None |
| 11 | Stock or stock options | X____None |
| 12 | Receipt of equipment, materials, drugs, medical writing, gifts or other services | X____None |
| 13 | Other financial or non-financial interests | X____None |

**Please place an "X" next to the following statement to indicate your agreement:**

**X___ I certify that I have answered every question and have not altered the wording of any of the questions on this form.**

### ICMJE DISCLOSURE FORM

**Date: 2021-03-03______________________________________________________________________________**
**Your Name: Hugo Sjöqvist________________________________________________________________________**
**Manuscript Title: Maternal Levels of Acute Phase Proteins in Early Pregnancy and Risk of Autism Spectrum Disorders in Offspring ___________________________________________________________________________**
**Manuscript number (if known):_________________________________________________________________**

**In the interest of transparency, we ask you to disclose all relationships/activities/interests listed below that are related to the content of your manuscript. "Related" means any relation with for-profit or not-for-profit third parties whose interests may be affected by the content of the manuscript. Disclosure represents a commitment to transparency and does not necessarily indicate a bias. If you are in doubt about whether to list a relationship/activity/interes it is preferable that you do so.**

**The following questions apply to the author's relationships/activities/interests as they relate to the current manuscript only.**

**The author's relationships/activities/interests should be defined broadly. For example, if your manuscript pertains to the epidemiology of hypertension, you should declare all relationships with manufacturers of antihypertensive medication, even if that medication is not mentioned in the manuscript.**

**In item #1 below, report all support for the work reported in this manuscript without time limit. For all other items, the time frame for disclosure is the past 36 months.**

| | | **Name all entities with whom you have this relationship or indicate none (add rows as needed)** | **Specifications/Comments (e.g., if payments were made to you or to your institution)** |
|---|---|---|---|
| | | **Time frame: Since the initial planning of the work** | |
| 1 | All support for the present manuscript (e.g., funding, provision of study materials, medical writing, article processing charges, etc.) **No time limit for this item.** | X____None | |
| | | **Time frame: past 36 months** | |
| 2 | Grants or contracts from any entity (if not indicated in item #1 above). | X____None | |
| 3 | Royalties or licenses | X____None | |
| 4 | Consulting fees | X____None | |
| 5 | Payment or honoraria for lectures, presentations, speakers bureaus, manuscript writing or educational events | X____None | |
| 6 | | X____None | |

| | | |
|---|---|---|
| | Payment for expert testimony | |
| 7 | Support for attending meetings and/or travel | X____None |
| 8 | Patents planned, issued or pending | X____None |
| 9 | Participation on a Data Safety Monitoring Board or Advisory Board | X____None |
| 10 | Leadership or fiduciary role in other board, society, committee or advocacy group, paid or unpaid | X____None |
| 11 | Stock or stock options | X____None |
| 12 | Receipt of equipment, materials, drugs, medical writing, gifts or other services | X____None |
| 13 | Other financial or non-financial interests | X____None |

**Please place an "X" next to the following statement to indicate your agreement:**

**X___ I certify that I have answered every question and have not altered the wording of any of the questions on this form.**

**ICMJE DISCLOSURE FORM**

**Date: 2021-03-03**____________________________________________________________
**Your Name: Christina Dalman**______________________________________________________
**Manuscript Title: Maternal Levels of Acute Phase Proteins in Early Pregnancy and Risk of Autism Spectrum Disorders in Offspring** __________________________________________________________
**Manuscript number (if known):**_______________________________________________________

**In the interest of transparency, we ask you to disclose all relationships/activities/interests listed below that are related to the content of your manuscript. "Related" means any relation with for-profit or not-for-profit third parties whose interests may be affected by the content of the manuscript. Disclosure represents a commitment to transparency and does not necessarily indicate a bias. If you are in doubt about whether to list a relationship/activity/interes it is preferable that you do so.**

**The following questions apply to the author's relationships/activities/interests as they relate to the <u>current manuscript only</u>.**

**The author's relationships/activities/interests should be <u>defined broadly</u>. For example, if your manuscript pertains to the epidemiology of hypertension, you should declare all relationships with manufacturers of antihypertensive medication, even if that medication is not mentioned in the manuscript.**

**In item #1 below, report all support for the work reported in this manuscript without time limit. For all other items, the time frame for disclosure is the past 36 months.**

| | | **Name all entities with whom you have this relationship or indicate none (add rows as needed)** | **Specifications/Comments (e.g., if payments were made to you or to your institution)** |
|---|---|---|---|
| | | **Time frame: Since the initial planning of the work** | |
| 1 | All support for the present manuscript (e.g., funding, provision of study materials, medical writing, article processing charges, etc.) **No time limit for this item.** | ____None | |
| | | Swedish Research Council | |
| | | **Time frame: past 36 months** | |
| 2 | Grants or contracts from any entity (if not indicated in item #1 above). | X____None | |
| 3 | Royalties or licenses | X____None | |
| 4 | Consulting fees | X____None | |
| 5 | Payment or honoraria for lectures, presentations, speakers bureaus, manuscript writing or educational events | X____None | |
| 6 | | X____None | |

| | | |
|---|---|---|
| | Payment for expert testimony | |
| 7 | Support for attending meetings and/or travel | X____None |
| 8 | Patents planned, issued or pending | X____None |
| 9 | Participation on a Data Safety Monitoring Board or Advisory Board | X____None |
| 10 | Leadership or fiduciary role in other board, society, committee or advocacy group, paid or unpaid | X____None |
| 11 | Stock or stock options | X____None |
| 12 | Receipt of equipment, materials, drugs, medical writing, gifts or other services | X____None |
| 13 | Other financial or non-financial interests | X____None |

**Please place an "X" next to the following statement to indicate your agreement:**

**X___ I certify that I have answered every question and have not altered the wording of any of the questions on this form.**
